# A novel neurodevelopmental syndrome caused by loss-of-function of the Zinc Finger Homeobox 3 (ZFHX3) gene

**DOI:** 10.1101/2023.05.22.23289895

**Authors:** María del Rocío Pérez Baca, Eva Z. Jacobs, Lies Vantomme, Pontus Leblanc, Elke Bogaert, Annelies Dheedene, Laurenz De Cock, Sadegheh Haghshenas, Aidin Foroutan, Michael A. Levy, Jennifer Kerkhof, Haley McConkey, Chun-An Chen, Nurit Assia Batzir, Xia Wang, Maria Palomares, Marieke Carels, ZFHX3 consortium, Bart Demaut, Bekim Sadikovic, Björn Menten, Bo Yuan, Sarah Vergult, Bert Callewaert

**Affiliations:** Center for Medical Genetics Ghent, Department of Biomolecular Medicine, Ghent University Hospital, Ghent, 9000, Belgium; Department of Biomolecular Medicine, Ghent University, Ghent, 9000, Belgium; Verspeeten Clinical Genome Centre, London Health Sciences Centre, London, ON N6A 5W9, Canada; Department of Pathology and Laboratory Medicine, Western University, London, ON N6A 3K7, Canada; Children’s Health Research Institute, Lawson Research Institute, London, ON, N6C 2R5, Canada; Baylor College of Medicine, Texas Children’s Hospital, TX 77030, USA; Schneider Children’s Medical Center of Israel, Petach Tikvah 4920235, Israel; INGEMM, Instituto de Genética Médica y Molecular, IdiPAZ, Hospital Universitario la Paz, Universidad Autónoma de Madrid (UAM), 28029 Madrid, Spain; VIB UGent Center for Inflammation Research, Department for Biomedical Molecular Biology, Ghent University, 9052, Belgium; Seattle Children’s Hospital, Seattle and Department of Laboratory Medicine and Pathology, University of Washington, WA 98105, USA

**Author notes:** corresponding authors Correspondence to: María del Rocío Pérez Baca & Sarah Vergult & Bert Callewaert Corneel Heymanslaan 10, Entrance 34, 9000 Ghent, Belgium. These authors contributed equally to this work (Sarah Vergult, Bert Callewaert). These authors contributed equally to this work (María del Rocío Pérez Baca, Eva Z. Jacobs). Appendix 1 (ZFHX3 consortium).

**Keywords:** neurodevelopmental disorder, ZFHX3, chromatin remodelling complex, mRNA polyadenylation and cleavage complex, *Drosophila melanogaster*

## Abstract

Neurodevelopmental disorders (NDDs) result from impaired development and functioning of the brain. Here, we identify loss-of-function variation in *ZFHX3* as a novel cause for syndromic intellectual disability (ID). ZFHX3, previously known as ATBF1, is a zinc-finger homeodomain transcription factor involved in multiple biological processes including cell differentiation and tumorigenesis.

Through international collaboration, we collected clinical and morphometric data (Face2Gene) of 41 individuals with protein truncating variants (PTVs) or (partial) deletions of *ZFHX3*. We used data mining, RNA and protein analysis to identify the subcellular localization and spatiotemporal expression of ZFHX3 in multiple *in vitro* models. We identified the DNA targets of ZFHX3 using ChIP seq. Immunoprecipitation followed by mass spectrometry indicated potential binding partners of endogenous ZFHX3 in neural stem cells that were subsequently confirmed by reversed co-immunoprecipitation and western blot. We evaluated a DNA methylation profile associated with *ZFHX3* haploinsufficiency using DNA methylation analysis on whole blood extracted DNA of six individuals with *ZFHX3* PTVs and four with a (partial) deletion of *ZFHX3*. A reversed genetic approach characterized the *ZFHX3* orthologue in *Drosophila melanogaster*.

Loss-of-function variation of *ZFHX3* consistently associates with (mild) ID and/or behavioural problems, postnatal growth retardation, feeding difficulties, and recognizable facial characteristics, including the rare occurrence of cleft palate.

Nuclear abundance of ZFHX3 increases during human brain development and neuronal differentiation in neural stem cells and SH-SY5Y cells, ZFHX3 interacts with the chromatin remodelling BRG1/Brm-associated factor complex and the cleavage and polyadenylation complex. In line with a role for chromatin remodelling, *ZFHX3* haploinsufficiency associates with a specific DNA methylation profile in leukocyte-derived DNA. The target genes of ZFHX3 are implicated in neuron and axon development.

In *Drosophila melanogaster*, z*fh2,* considered to be the *ZFHX3* orthologue, is expressed in the third instar larval brain. Ubiquitous and neuron-specific knockdown of zfh2 results in adult lethality underscoring a key role for zfh2 in development and neurodevelopment.

Interestingly, ectopic expression of zfh2 as well as ZFHX3 in the developing wing disc results in a thoracic cleft phenotype.

Collectively, our data shows that loss-of-function variants in *ZFHX3* are a cause of syndromic ID, that associates with a specific DNA methylation profile. Furthermore, we show that ZFHX3 participates in chromatin remodelling and mRNA processing.

## Introduction

Neurodevelopmental disorders (NDDs) are a heterogeneous group of early onset conditions that impact development and functioning of the central nervous system (CNS) affecting approximately 2-5% of children worldwide. The clinical and genetic heterogeneity of NDDs make it challenging to find a molecular diagnosis for individual cases^1^. Although recent technological improvements led to a significant increase in diagnostic yield and the identification of several new NDD associated genes, many associated genes remain to be discovered.^2^ Here, we identify loss-of-function defects in *ZFHX3* (Zinc Finger Homeobox 3) in a novel NDD.

*ZFHX3*, previously known as AT motif-binding transcription factor 1 (*ATBF1*), encodes one of the largest transcription factors (404 kDa) containing 23 zinc finger motifs and four homeodomains. It was originally identified as a transcriptional suppressor that binds an AT-rich motif in the alfa-fetoprotein enhancer in human hepatoma cells^3^. Subsequently, ZFHX3 was found to function in multiple biological processes including mammary gland development^4, 5^, myogenic differentiation^6^, and pathological processes as tumorigenesis^7–9^ and atrial fibrillation^10–12^. In mice, expression of *Zfhx3* is high during embryonic stages of CNS development but ceases upon brain maturation^13^. Nevertheless, expression remains enriched in the adult suprachiasmatic nucleus where it is involved in circadian function^14^.

*Zfhx3* expression also continues in cortical excitatory neurons and appears in late differentiating nonoverlapping neuron populations^15^. These results correlate with previous work from Moreau et al., showing that Zfhx3 belongs to the Pbx3 neuron cluster, an early cortical excitatory neuron subtype^16^. *Zfhx3* is further selectively expressed in immature dopamine receptor expressing medium spiny neurons within mice striatum^17^. In further support of a role in neuronal differentiation, *Zfhx3* expression increases upon neuronal differentiation of pluripotent mouse embryonal carcinoma P19 cells^18, 19^ Similarly, in the zebrafish, *zfhx3* expression coincides with regions of neurogenesis at critical developmental time points^20^.

*ZFHX3* is predicted to be intolerant to protein truncating variants (PTVs), reflected by the **pLI-score (**probability of being loss-of-function intolerant) of 1 and a constraint score of observed/expected (oe) of 0.08 in the gnomAD database^21^. Moreover, it belongs to the top 5% of genes implicated in monoallelic autism spectrum disorder (ASD) and/or developmental delay (DD) according to the NDD risk gene prediction tool mantis-ml (https://nddgenes.com)^22^ and was suggested as an NDD candidate gene susceptible to *de novo* likely gene disruptive variation by the shallow neural network developed by Chow & Hormozdiari^23^.

In this study, we define the phenotypic spectrum in an extensive cohort of 42 individuals with loss-of-function variants (18 (partial) deletions and 24 truncating variants) affecting the *ZFHX3* gene. We report the spatio-temporal and subcellular expression patterns of ZFHX3 during human brain and neuronal stem cell development and show that its DNA target genes are implicated in neuron and axon development. Furthermore, ZFHX3 contributes to pre-messenger RNA (pre-mRNA) processing, chromatin remodeling and cytoskeleton reorganization. We define expression of *zfh2*, the orthologue of ZFHX3 in *Drosophila melanogaster,* in the larvae brain and show that overall knockdown leads to lethality and ZFHX3 ectopic expression phenocopies zfh2 ectopic expression. Overall, our data identify ZFHX3 as an important regulator in neuronal differentiation.

## Materials and methods

### Patient recruitment

Data sharing through GeneMatcher^24^, DECIPHER^25^ or personal communication identified 42 probands across 42 institutions in 16 countries with truncating variants in *ZFHX3* (n=24) and (micro)deletions affecting *ZFHX3* (n=18). Inheritance of *ZFHX3* variants was *de novo* (n=32), inherited from a (mildly) affected parent (n=2), or unknown (n=8). All affected probands were investigated by their referring physicians and all genetic analyses were performed in a diagnostic setting. An overview of the clinical presentation of all probands and family members is given in Supplementary Tables 1 and 2. A summary of the clinical characteristics is presented in Table 1. Legal guardians of affected probands gave informed consent for genomic investigations and publication of pseudonymized data following the rules of the Helsinki declaration and according to EC2019/1430 of Ghent University Hospital and/or local ethical committees of the referring centers and the Western University Research Ethics Board (REB 106302).

**Table 1.**
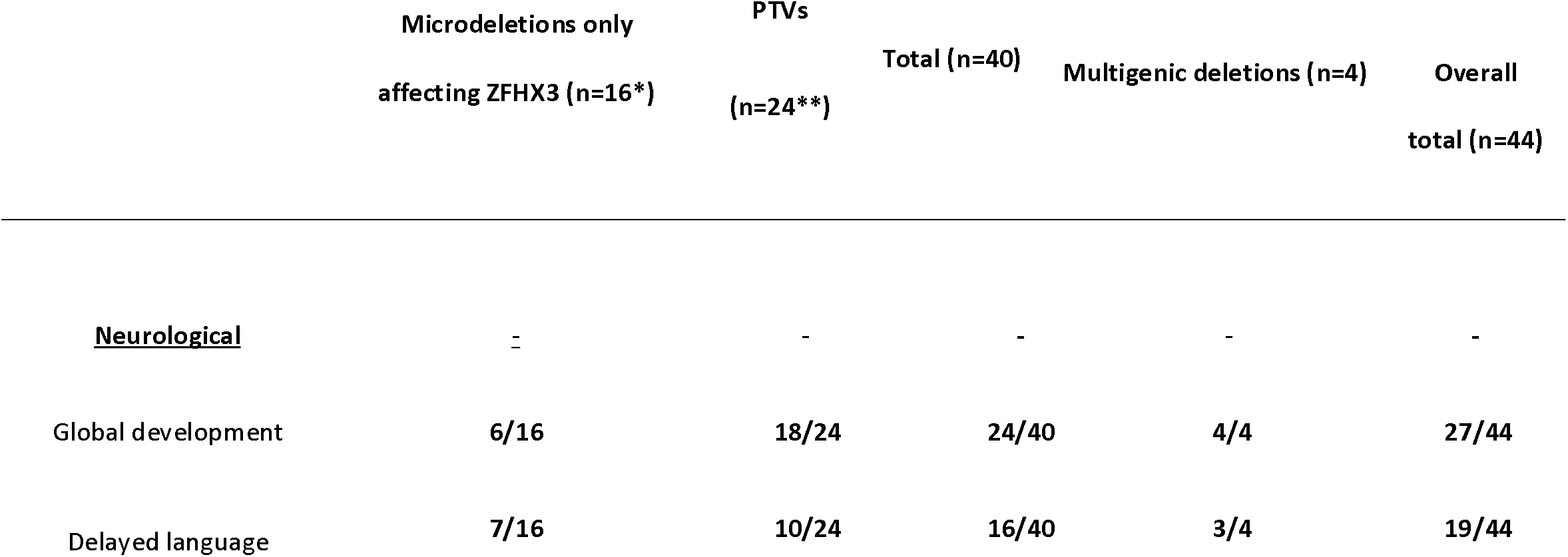
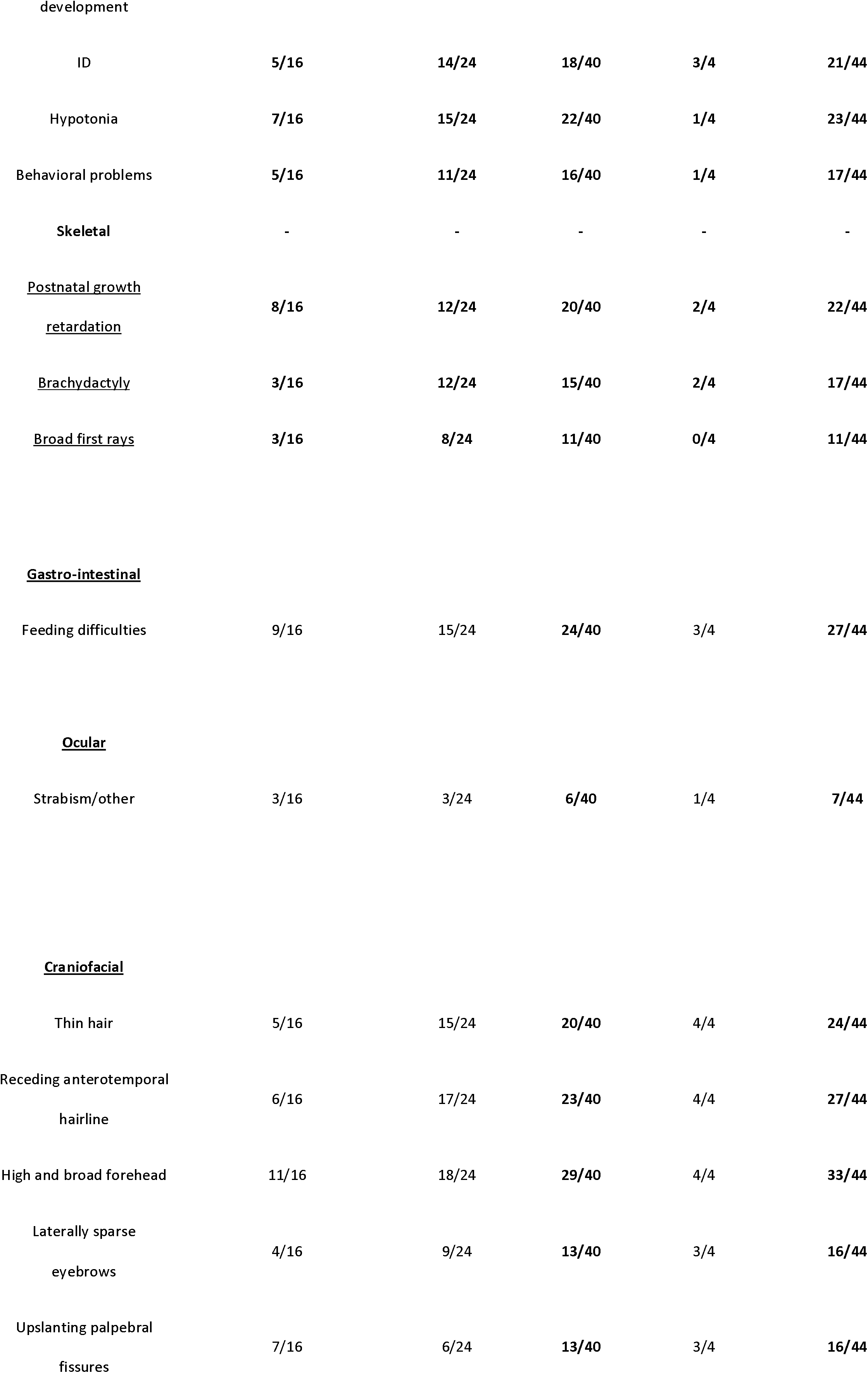
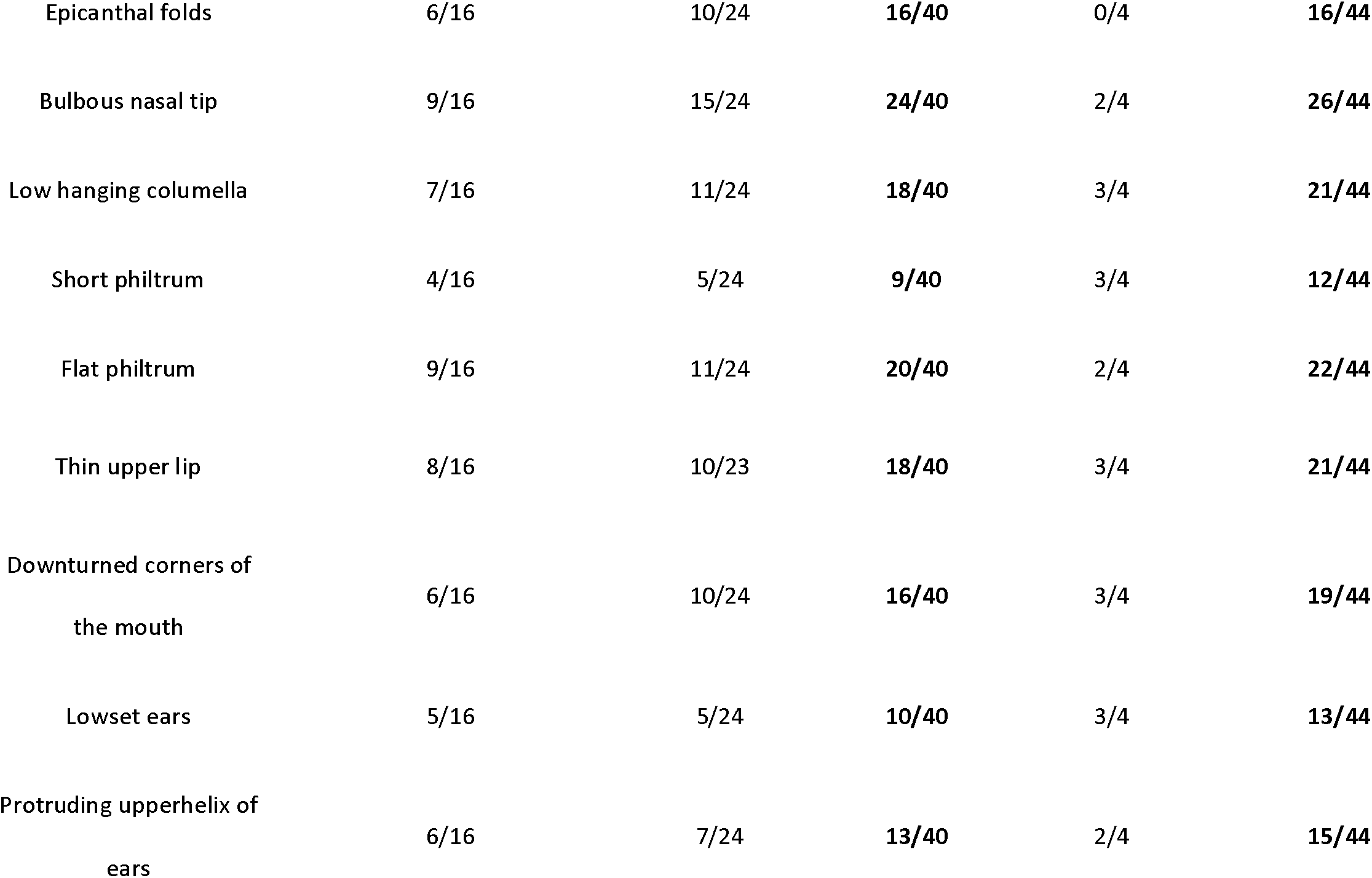
Clinical characteristics of 38 probands and 6 affected family members with a microdeletion or a protein truncating variant (PTV) affecting ZFHX3. *x PTV probands & x affected family members, **x microdeletions probands & x affected family members

### Facial features analysis

Frontal facial images of 17 individuals with either truncating variants in *ZFHX3* or microdeletions only affecting *ZFHX3* were uploaded to the Face2Gene Research app (FDNA Inc., Boston, MA, USA), de-identified and analyzed through the use of the DeepGestalt image analysis technology, a deep-learning computer algorithm^26^. The resulting artificial composite image was compared to that of a control cohort matched by age, sex, and ethnicity in the same way as described by Mak et al. 2021^27^. This results in a Score Distribution and a receiver operating characteristic curve (ROC curve).

### Cell culture

#### SH-SY5Y and HEK293T

SH-SY5Y and HEK293T cells were cultured in RPMI 1640 medium (52400041, Gibco^TM^) supplemented with 10% fetal bovine serum (FBS), 2 mM L-Glutamine and 100 IU/ml penicillin/streptavidin at 37°C in a 5% CO2 humid atmosphere.

#### Human embryonic stem cells (hESCs)

Naïve human embryonic stem cells (hESCs) cultured on CF-1 irradiated Mouse Embryonic Fibroblasts (MEFs) (A34180, Gibco^TM^) were kindly provided by the group of Björn Heindryckx, Ghent University (Duggal et al., 2015)^28^ and converted to feeder-free naïve-like human embryonic stem cells with RSeT Feeder-Free medium (STEMCELL^TM^ technologies) according to manufacturer’s instructions with an adapted in-house protocol (derived from Aalders, et al. 2019)^29^ described in the Supplementary Material.

#### Neural stem cells (NSCs)

The BJ fibroblast cell line from human foreskin (#CRL-2522, ATCC) was reprogrammed to human induced pluripotent stem cells (iPSCs) using the CytoTune-iPS 2.0 Sendai Reprogramming Kit (Invitrogen). Pluripotency of iPSC clones was validated with immunostaining for pluripotent markers and an embryoid body assay. These iPSCs were differentiated to NSCs using PSC Neural Induction Medium (Gibco^TM^, ThermoFisher). These NSCs were kindly provided by the group of Peter Ponsaerts, University of Antwerp. The NSC line was further expanded in Neural Expansion Medium (Gibco^TM^, ThermoFisher) on coated cultured vessels with Geltrex® LDEV-Free hESC-Qualified Reduced Growth Factor Basement Membrane Matrix (Gibco^TM^, ThermoFisher), according to the manufacturer’s instructions. The neural stem cell line was cultured at 37°C in a 5% CO2 humid atmosphere.

#### Neural differentiation using BrainPhys media (Stemcelltechnologies)

Neural stem cells were cultured in BrainPhys^TM^ Neuronal Medium (STEMCELL Technologies), supplemented with NeuroCultTM SM1 Neuronal Supplement (STEMCELL Technologies), N2 Supplement-A (StemCell Technologies), recombinant Human Brain Derived Neurotrophic Factor (BDNF, PeproTech, 20 ng/mL), recombinant Human Glial-Derived Neurotrophic Factor (GDNF, PeproTech, 20 ng/mL), dibutyryl cAMP (1 mM, Sigma), and ascorbic acid (200 nM, Sigma). Half of the total volume was replaced at every 2–3 days.

### Immunofluorescence

SH-SY5Y and NSCs were cultured onto glass coverslips in 12-well plates and fixed with 3.7% formaldehyde, for 20 min at room temperature. Firstly for the NSCs, a specific coating (Geltrex™ LDEV-Free, hESC-Qualified, Reduced Growth Factor Basement Membrane Matrix, Gibco^TM^, Thermofischer) was embedded into the coverslips. The cells were further stained following the protocol extensively described in the Supplementary Material. Imaging was performed by a Zeiss Axio Observer Z1 fluorescent microscope (Carl Zeiss, Australia). The magnification of the images was 100x.

### Expression profiling

RNA of cultured cells was isolated using the Direct-zol™ RNA MiniPrep Kit (Zymo Research) according to manufacturer’s instructions. NanoDrop (Thermofischer) was used to determine the concentration. cDNA synthesis was performed using the iScript Advanced cDNA synthesis kit (BioRad) according to manufacturer’s instructions. Subsequently, RT-qPCR was performed with a PCR mix containing 5 ng cDNA, 2.5 µl of SsoAdvanced SYBR qPCR mix (BioRad) and 0.25 µl of forward and reverse primers (250 µM concentration, Integrated DNA Technologies). The RT-qPCR was performed on a LC-480 device (Roche) and gene expression levels were analyzed using the qBase+ software 3.2 (Biogazelle). The RT-qPCR primers used in this study can be found in Supplemental Table 11.

### Immunoprecipitation followed by mass-spectrometry (IP-MS)

2x10^7^ cells (i.e. SH-SY5Y and NSCs) were resuspended in 1000 μl freshly made RIPA buffer and rotated for 1h at 4°C to lyse the cells. Following centrifugation of the cells for 10 min at 8000g, 2 μg antibody (anti-ZFHX3, #PD010, MBL; anti-IgG, ab2410, Abcam) was added to 500 µl supernatant and rotated for 4h at 4°C. The immunoprecipitation was performed with protein A Ultralink resin beads (Thermo Scientific) following the protocol found in the Supplementary Material. For each cell line, three biological replicates were included.

### Expression Constructs, Transfection, Cell harvesting & Protein extraction

The ZFHX3-FLAG plasmid was ordered from AddGene (pcDNA3.1-FLAG-ATFB1, (#40927). *SMARCB1* and *CPSF2* constructs were ordered from Genecopoeia as shuttle vectors. Subsequently, Gateway cloning (Gateway Technology, Gateway LR reaction, Invitrogen) was performed according to the manufacturer’s instructions to insert the shuttle vectors’ Open Reading Frame (ORF) (*SMARCB1* or *CPSF2*) into a destination vector pMH-HA N-term tag vector. A N-term 3xHA-tag followed the ORF in the new designed SMARCB1-3xHA or CPSF2-3xHA expression vector. All constructs were Sanger sequenced (GATC sequencing, Eurofins Genomics) bidirectionally for their entire open reading frame. The vector maps can be found in Supplemental Figure 8.

To confirm protein-protein interaction via co-IP (see below), the ZFHX3-FLAG construct was co-transfected with the SMARCB1-3xHA or the CPSF2-3xHA expression vector in HEK293T cells. As control conditions, we performed single transfections of ZFHX3-FLAG, SMARCB1-3xHA, or CPSF2-3xHA alone. Transfection complexes were prepared according to the Lipofectamine 3000 solution manufacturer’s instructions (Supplementary Material).

Upon 48 hours incubation time, 2.5*10^6^ cells were harvested per condition and lysed in freshly made RIPA buffer for protein extraction. Following centrifugation (10min, 4°C, 8000g), proteins were extracted, and the concentration was determined using the BCA protein assay kit (Thermo Fisher). Overexpression of the protein of interest was confirmed by Western Blot (Supplementary Material).

### Co-immunoprecipitation (co-IP)

Immunoprecipitation was performed according to the Dynabeads^TM^ Protein G (Invitrogen) manufacturer’s instructions. 300μl lysate was added to the antibody beads complex and incubated overnight at 4°C. Protein complexes were eluted with 20 μl glycine buffer (pH 2.8) and determined using Western Blot. As a negative control for the co-immunoprecipitation (co-IP), 300μl per cell lysate was added to Dynabeads^TM^ protein G, coupled to non-specific IgG antibodies.

The anti-HA.11 Epitope Tag antibody (1/1000, Clone 16B12, Biolegend) was used to enrich the capture of the SMARCB1-HA tagged overexpressed protein and the CPSF2-HA tagged overexpressed protein within the co-IP.

### Western Blot

Western blot was performed according to an in-house protocol described in the Supplementary Material.

### DNA methylation analysis

The workflow used to establish whether a specific DNA methylation profile is found within our LoF cohort in comparison with other neurodevelopmental syndromes, is described in detail in Aref-Eshghi et al.^30^. Further description can be found in the Supplementary Material.

### ChIP-sequencing and data analysis

ChIP-seq for ZFHX3 has been performed on 10 million NSCs (with endogenous ZFHX3 expression) and 10 million NSCs with overexpressed ZFHX3 expression. These experiments were conducted by the Diagenode ChIP-seq/ChIP-qPCR Profiling service (Diagenode Cat# G02010000). Material and methods are described in the Supplementary Material.

Using ChIPSeeker^31^, the ChIP peaks called in both the sample with endogenous ZFHX3 expression and the sample with ectopic ZFHX3 expression, were annotated to the nearest gene based on the distance of the peak to the nearest transcription start site (TSS) with a max distance cut-off of 1kb using the annotations from the TxDb.Hsapiens.UCSC.hg38.knownGene package. To identify predominant biological processes (BP), we performed functional enrichment analysis using the BP gene ontology (GO) terms of the annotated genes. The top20 of enriched GO terms (q-value<0.05) is visualized as a dotplot in Figure 5C. In Figure 5D, the Kyoto Encyclopedia of Genes and Genomes (KEGG) analysis provides the annotation of biological pathways. The top10 of enriched pathways (q-value<0.05) is visualized as a dotplot.

### Mining of public data

#### Human expression datasets

Expression levels in terms of transcripts per million (TPM) were retrieved from Cardoso-Moreira et al.^32^ We examined the expression of *ZFHX3* in seven human tissues (i.e. brain, cerebellum, heart, kidney, liver, ovary and testis) and developmental stages (see Supplemental Figure 2B). Furthermore, for the developmental data of these seven human organs, Spearman’s correlations were computed between expression values of *ZFHX3* and expression values of all other genes. Subsequently these data were ranked based on the Rho value (> 0.8) and a p-value <0.01. This ranked list was used to perform a preranked GSEA (https://www.gsea-msigdb.org/gsea/index.jsp) with the following settings: Gene Set “C5 GO:MF ”, number of permutations: 1000, min/max number of genes per gene set: 15/200. The GO terms of the top20 enriched gene sets (all with normalized enrichment score > 2 and FDR <0.10) of this GSEA preranked analysis were subsequently visualized.

In addition, the ZFHX3 expression levels in terms of TPM (transcripts per million) were retrieved from GTEX (https://www.gtexportal.org/home/). We examined the expression of ZFHX3 and its different isoforms in 54 non-diseased adult tissues (see Supplemental Figure 2A and Supplemental Figure 4). Furthermore, the ZFHX3 expression levels in terms of RPKM (reads per kilobase million) from six brain regions during neurodevelopment and adulthood were retrieved from Brainspan (http://www.brainspan.org, see Figure 2C). Moreover, single cell ZFHX3 expression data during neuronal differentiation from stem cells to neural progenitor cells and mature neurons on the one hand and to one- or five-months undirected cerebral organoids on the other hand, was respectively retrieved from https://bioinf.eva.mpg.de/shiny/sample-apps/scApeX/^33^ (Supplemental Figure 2C) and https://shcheglovitov.shinyapps.io/u_brain_browser/^34^ (Supplemental Figure 3).

#### *Drosophila* zfh2 ChIP-seq dataset

Kudron et al. published a catalog of regulatory sites (The ModERN Resource) containing several transcription factor binding sites in *Drosophila melanogaster* and *Caenorhabditis elegans*^35^. The chromatin immunoprecipitation (ChIP) sequencing data from the transgenic flies expressing *zfh2*-eGFP fusion protein from this resource, is available through the Encode Project database (https://doi.org/doi:10.17989/ENCSR259WOE). Using ChIPSeeker^31^, the ChIP peaks were annotated to the nearest gene based on the distance of the peak to the nearest transcription start site (TSS) with a max distance cut-off of 1kb using the annotations from the TxDb.Dmelanogaster.UCSC.dm6.ensGene package. From the functional enrichment analysis using the BP GO terms, the top20 of enriched GO terms (q-value<0.05) is visualized as a dotplot in Figure 6C.

### Drosophila stocks and the generation of the transgenic lines

The fly lines used in this study were obtained from the Bloomington Drosophila Stock Center (BDSC), Vienna Drosophila Resource Center (VDRC) or generated by Genetivision. The fly lines are described in the Supplementary Material. All flies were cultured at 25°C on standard cornmeal and molasses medium in plastic vials, unless otherwise noted.

### Immunofluorescence and confocal microscopy for larval brains

Larval brains from all genotypes were dissected in PBS, and fixed in 4% paraformaldehyde in PBS, for 10 min at room temperature. The brains were further immunostained following the protocol described in the Supplementary material. The imaging was performed with a Leica SP8 confocal microscope and processed using ImageJ.

### Bright field Imaging of Flies

Images of the *Drosophila* were taken using a Leica DFC 450C camera mounted on a microscope Leica Z16 APO using LAS V4.12 software. The images were processed using a LAS V4.12 software to obtain stack images.

### Offspring assay

Crosses for adult offspring frequencies were performed at 25° and 29°C. For each *Drosophila* cross the collected offspring was divided by sex and the genotypes were counted according to the balancers. The offspring ratio was determined by dividing counted offspring by expected offspring. Lethality was attributed to conditions where no flies of the expected phenotype were observed in a minimum total offspring of 100 flies.

## Data availability

Public data was used from: https://apps.kaessmannlab.org/evodevoapp/

https://www.gtexportal.org/home/

http://www.brainspan.org

https://bioinf.eva.mpg.de/shiny/sample-apps/scApeX/

https://shcheglovitov.shinyapps.io/u_brain_browser/

https://gnomad.broadinstitute.org/about

https://doi.org/doi:10.17989/ENCSR259WOE

The analyzed IP-MS data and the overlapping peaks between endogenous and ectopic ZFHX3 ChIP-seq is available in the Supplementary Material.

Some of the DNA methylation datasets used in this study are publicly available and may be obtained from gene expression omnibus (GEO) using the following accession numbers. GEO: GSE116992, GSE66552, GSE74432, GSE97362, GSE116300, GSE95040, GSE 104451, GSE125367, GSE55491, GSE108423, GSE116300, GSE 89353, GSE52588, GSE42861, GSE85210, GSE87571, GSE87648, GSE99863, and GSE35069. These include DNA methylation data from patients with Kabuki syndrome, Sotos syndrome, CHARGE syndrome, immunodeficiency-centromeric instability-facial anomalies (ICF) syndrome, Williams-Beuren syndrome, Chr7q11.23 duplication syndrome, BAFopathies, Down syndrome, a large cohort of unresolved subjects with developmental delays and congenital abnormalities, and also several large cohorts of DNA methylation data from the general population. Remaining methylation data is not publicly available due to institutional and ethics restrictions.

## Results

### Individuals carrying a *ZFHX3* deletion or protein truncating variant present with a NDD phenotype

#### Molecular characterization

Eighteen probands harbor a microdeletion in chromosome band 16q22.2-16q22.3 ranging in size from 53.04 kb to 8.51 Mb (probands 1-18; Supplemental Table 1) resulting in either partial or complete deletion of *ZFHX3* (Figure 1A). The deletion in probands 4-14 and 16-18 only affects one protein-coding gene, i.e. *ZFHX3.* In probands 4-8, 15, 17 and 18, the neighboring long-noncoding RNA (lncRNA) genes *LINC01572*, *LINC01568* and *HCCAT5* are also affected. The function of these lncRNAs is hitherto unknown. The deletion in probands 1, 2, 3 and 15 involves additional genes (see Supplemental Table 9). Inheritance could be determined for fourteen deletions: thirteen occurred *de novo* (probands 1-6; 12-18) and one was inherited from the similarly affected father (proband 10; also present in similarly affected sister). Parental segregation was incomplete for proband 7, 8, 9 & 11; the deletion in proband 11 is absent in the mother, but could not be investigated in the father.

**Figure 1:**
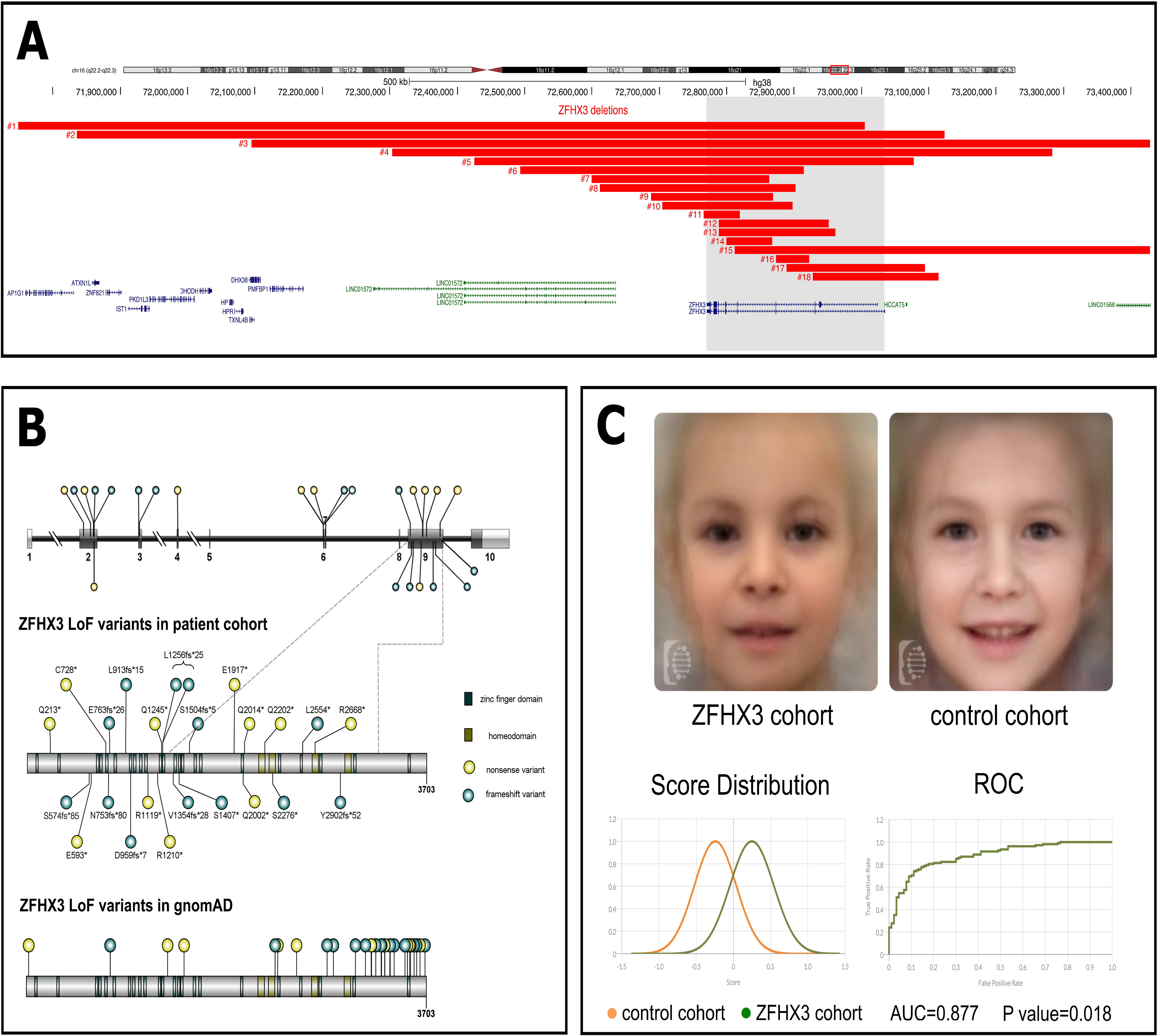
Overview of *ZFHX3* aberrations in 42 probands. A. (Micro)deletions identified in probands 1-18 (#) are represented by the red bars, all affecting the *ZFHX3* gene (grey box). RefSeq coding and non-coding genes are indicated respectively in blue and green. Genomic positions are according to hg38, *ZFHX3* is located on the reverse strand. **B**. Down & Left: Nonsense and frameshift variants in *ZFHX3* are identified in probands 19-42. Gene and protein structure of the canonical transcript (NM_006885.3; NP_008816.3) are shown in the top and middle part, respectively. The majority of PTVs are located in exon 9. **Top**: coding exons are indicated as dark grey boxes, while UTR regions are indicated with a light grey bar. **Middle:** LoF variants are present in the ZFHX3 patient cohort and located in different domains of the ZFHX3 protein. **Bottom:** LoF variants present in gnomAD are enriched at the C-terminus, i.e. 18 of the 35 reported variants are located in exon 10. C. Down & Right: Clinical presentation of individuals with *ZFHX3* aberration. Top: De-identified face mask of images from 17 individuals with either a microdeletion or truncating variant affecting *ZFHX3 (left)* and matched healthy individuals (right). Note a high and broad forehead, laterally sparse eyebrows, upslanted palpebral fissures, a low-hanging columella, flat philtrum, a relatively short midface, and a thin upper lip. Bottom: Score distribution plots of the ZFHX3 cohort (green) and the control cohort (orange) and the receiver operating characteristic (ROC) curve to determine the capacity of Face2Gene to identify *ZFHX3* cases against controls.

Twenty-four probands harbor a PTV in *ZFHX3* of which 19 PTVs were confirmed *de novo* (probands 20-29; 31-36; 38-40), one nonsense *ZFHX3* variant segregates with the phenotype (proband 38 and four affected family members) (Figure 1B; Supplementary Table 2). Inheritance could not be determined for the frameshift variants in probands 19, 30, 41 and 42. However, the same frameshift variant identified in proband 30 was also observed *de novo* in proband 31.

All variants are absent from the public variant database gnomAD (release v3.1.1) and are predicted to result in protein truncation or to initiate nonsense mediated decay (NMD). Furthermore, all eleven nonsense variants have a CADD-score greater than 36 (Supplemental Table 2), indicating they are all predicted to be within the 0.1% most deleterious possible substitutions in the human genome. Nine missense variants present in gnomAD affect the same codon as the truncating variant in probands 21, 36, 37 and 42 (Supplemental Table 3), suggesting these missense variations do not result in loss of function (LoF) outcome. Missense variation seems well tolerated based on the z-score of -1.23, while the pLI score of 1.0 and LOEF score (loss-of-function observed/expected upper bound fraction) of 0.14 clearly indicate intolerance of LoF variation. Almost half of the truncating variants in the patient cohort are located in the large and penultimate exon 9 (11/24). Furthermore, all PTVs present in our cohort are at least 50 base pairs upstream of the last exon-exon boundary (Figure 1B). No truncating variants are observed at the C-terminus in the patient cohort, while the predicted LoF variants present in gnomAD are enriched at the C-terminus (i.e. exon 10) suggestive for NMD escape (Figure 1B).

#### Phenotypic characterization

Probands 1, 2, 3 and 15 harboring multigenic deletions were excluded for the characterization of the *ZFHX3*-associated phenotype to avoid a possible bias due to haploinsufficiency of other genes (Table 1, Supplemental Tables 1, 2 & 9). In addition, besides neurodevelopmental delay, no detailed clinical information could be obtained for probands 8, 9, 14, 19 & 25. Therefore, the phenotypic characterization is based on 33 probands and six (similarly) affected family members. Early neurological development is characterized by global neurodevelopmental delay (24/39), delayed language (17/39) and/or motor (26/39) development, the latter often preceded by hypotonia (22/39). Intellectual capacities vary from normal with or without some learning problems (n=14) to mild and moderate intellectual impairment (n=19). Behavioral problems (16/39) are variable. Autism spectrum disorder (ASD) was noted in nine individuals but is the sole neurological manifestation in proband 27, 38 and 39. Seizures were reported in two individuals (probands 28 and 35). Frequently associated features are postnatal growth retardation (20/39) and feeding difficulties (24/39) due to oropharyngeal dysphagia, eosinophilic esophagitis (n=2), or food allergies. Three individuals needed a gastrostomy tube. Hands and feet may be small and show brachydactyly (mostly of the distal phalanges, n=15), clinodactyly (mostly of the 4^th^ or 5^th^ rays, n=6), tapering fingers (n=3), and/or abnormal dermal creases (n=6). Other skeletal features include scoliosis and kyphosis (n=3), pectus deformity (n=3), hip dysplasia (n=3), and osteoporosis/fractures. Five individuals reported musculoskeletal pains, joint laxity, and/or fatigue. Cardiovascular features include septal defects in four patients, and two patients showed arterial stenoses. Sporadic urogenital defects include pyeloureteral junction stenosis (proband 4), vesicoureteral reflux (proband 27), renal cysts (proband 26), and cryptorchidism (probands 15 and 38). Hearing is reportedly normal, but several probands received transtympanic drains (n=7/39)

Facial gestalt comprises a receding anterotemperal hairline (23/39), high and broad forehead (29/39), bulbous nasal tip (24/39), low hanging columella (18/39), flat philtrum (17/39), thin upper lip (18/39) & thin vermillion border (18/39). Less common facial features include protruding ears (13/39), laterally sparse eyebrows (13/39), upslanted palpebral fissures (13/39), epicanthal folds (16/39), a short philtrum (9/39) and downturned corners of the mouth (16/39). Ocular abnormalities (ptosis, strabismus, nystagmus, tear duct aplasia, coloboma) are nonspecific and uncommon. Figure 1C shows a facial composite image of 17 patients with a *ZFHX3* aberration, compared to a healthy cohort matched for age and ethnicity. The ROC curves in Figure 1C show that Face2Gene was able to accurately categorize the ZFHX3 cohort vs controls 87.7% of the time (Area Under the Curve AUC=0.877) with a p-value of 0.018. The score distribution shows that there is a subtle but clear difference between the facial features of our ZFHX3 cohort in comparison with those of the control cohort.

The individuals with multiple deleted genes share most of the clinical characteristics but show a more severe developmental phenotype (Table 1). Gender (22 males and 20 females) is equally distributed and does not influence the clinical presentation. Age ranges from 10 months to 63 years. Overall, no clear differences could be observed between individuals with a microdeletion only affecting *ZFHX3* and a truncating variant in *ZFHX3*. Detailed clinical characteristics per individual can be found in Supplemental Table 1 (Deletions) & Supplemental Table 2 (PTVs). Pictures of front and side profiles as well as hands and digits of several probands (and affected family members) can be found in Supplemental Figure 1.

### ZFHX3 expression increases during human neurodevelopment

Publicly available RNA-seq data from 54 non-disease tissue sites, derived from mostly older donors (GTEx, release v8), indicates abundant *ZFHX3* expression throughout the human adult body, with highest expression in arteries (Supplemental Figure 2A). Expression in various adult brain regions is relatively low. Evaluation of ZFHX3 expression profiles across 7 organs and 22 developmental stages^32^, shows highest expression of ZFHX3 during the prenatal period, especially in the human brain (Supplemental Figure 2B). This is also confirmed by expression data from Brainspan, showing highest ZFHX3 expression in the brain prenatally, especially in the striatum, dorsal thalamus and mediodorsal nucleus of the thalamus (Figure 2B). This prenatally enriched expression pattern indicates/suggests a role during early (brain) development.

We observed increased expression of ZFHX3 during *in vitro* neural differentiation starting from human embryonic stem cells (hESCs), with the highest expression in early developed neurons (Figure 2A). Kanton et al.^33^ confirmed our observation with single cell RNA-seq data during cerebral organoid differentiation starting from pluripotency (Supplemental Figure 2C). Furthermore, single-cell sequencing data generated from single neural rosettes (SNR) derived organoids by the Shcheglovitov lab, reveals that *ZFHX3* is particularly expressed in 1 month old organoids in inhibitory neurons (Supplemental Figure 3)^34^.

**Figure 2:**
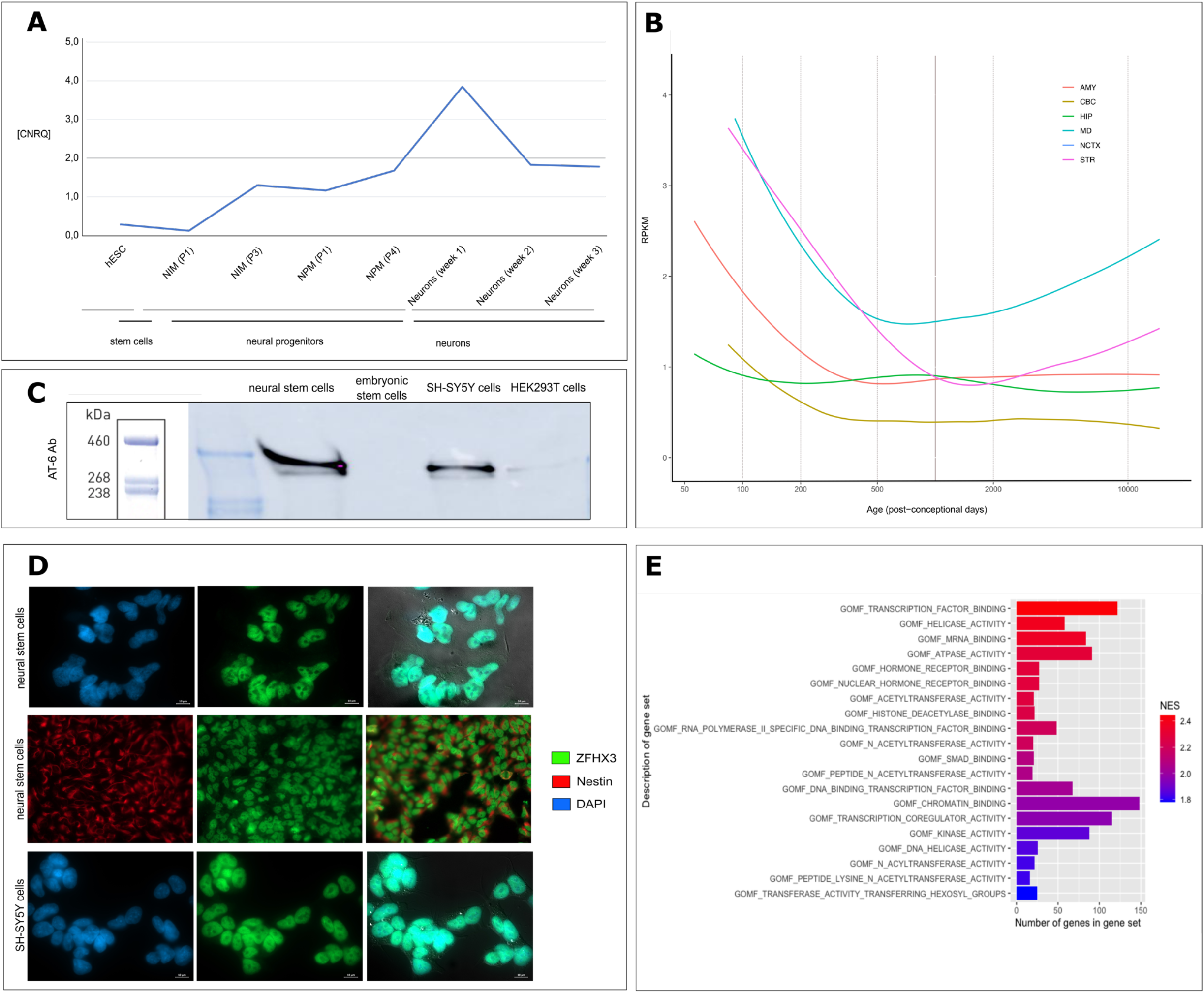
ZFHX3 is increasingly expressed during neural differentiation and localizes to the nucleus. **A. ZFHX3 expression during *in vitro* neural differentiation.** RNA was sampled at three stages: hESCs, neural progenitor cells and neurons. An increasing expression is observed upon differentiation. CNRQ: Calibrated Normalized Relative Quantity. NIM: STEMdiff Neural Induction Medium. NPM: STEMdiff Neural Progenitor Medium. P: passage. **B. Expression of ZFHX3 in developing human brain.** ZFHX3 expression in the developing human brain (normalized RPKM data) showing higher expression during early prenatal development followed by decreased expression upon brain maturation. Data obtained from BrainSpan: http://www.brainspan.org. Abbreviations: AMY, amygdala; CBC, cerebellum; HIP, hippocampus; MD: medial dorsal nucleus of the thalamus; NCTX: neocortex; STR: striatum. The vertical line indicates time of birth. **C. Endogenous ZFHX3 (404 kDa) expression in NSCs, hESCs, SH-SY5Y and HEK293T** as detected with Western Blot using the AT-6 antibody. The highest protein expression is observed in the immature neuronal cell lines SH-SY5Y and NSCs. **D. ZFHX3 is expressed in the nucleus of SH-SY5Y cells and NSCs.** Indirect immunofluorescence staining for ZFHX3 (SH-SY5Y and NSCs) and nestin (cytoplasmic neuronal marker, NSCs) was performed. DNA in the nucleus was counterstained with Hoechst dye. Images were merged to observe the contrast between the compartments. Scale bar: 10μm.; hESC: human embryonic stem cells; NSC: neural stem cells. **E. Molecular functions of the top 20 positive correlated gene sets in the data by Cardoso et al, 2019**^32^. All have an FDR <0.01 and a normalized enrichment score (NES) >1.8. Gene sets are ranked based on NES-value.

To screen for potential functional and regulatory interactions of ZFHX3, we performed a correlation analysis between expression of ZFHX3 and all other genes across the seven organs and 22 developmental stages assessed by Cardoso-Moreira et al. Supplemental Table 4 contains the rho and p-values with significant correlation (p-value <0.05). Top correlated genes appear to play a role in transcription factor binding, ac(et)yltransferase activity, kinase activity, chromatin binding and mRNA binding (Figure 2E).

### ZFHX3 has two isoforms and is expressed in the nucleus of different neuronal cell lines

There are two main ZFHX3 isoforms, previously called ATBF1-A (NM_006885; ENST00000268489) and ATBF1-B (NM_001164766; ENST00000397992), generated by alternative splicing and promotor usage. ATBF1-A encodes a 404-kDa protein and contains 23 zinc finger and four homeodomains. ATBF1-B (306-kDa) contains the same homeodomains, but lacks five zinc finger domains due to the absence of 920 amino acids at the N-terminus^18^. In 18/24 of the cases with a *ZFHX3* PTV, the aberration affects both the short and long isoform. The variants identified in probands 19-24 only affect the longest isoform.

According to GTEx data, the 306-kDa isoform shows higher expression at mRNA level in all adult tissues (Supplemental Figure 4). Nevertheless, the public data we leveraged indicate that ZFHX3 is particularly expressed in the prenatal brain and that expression in the adult brain is much lower^32^ (Figure 2B & Supplemental Figure 2B). We therefore evaluated the expression of ZFHX3 in human embryonic stem cells (hESC), two immature neuronal-like cell lines (SH-SY5Y a neuroblastoma cell line, often used as neuronal *in vitro* model) and human neural stem cells (NSCs)), and a non-neuronal cell line (human embryonic kidney cells (HEK293T)) at protein level, showing that the long 404-kDa isoform is mainly expressed in immature neuronal-like cell lines (i.e. SH-SY5Y & NSCs) (Figure 2C). We then investigated the location of ZFHX3 within these two cell lines to correlate subcellular expression with its potential functions. Immunocytochemistry for ZFHX3 in these two cell lines shows nuclear expression correlating with its function as a transcriptional factor (Figure 2E & Supplemental Figure 5).

### ZFHX3 interacts with members of the BAF and CP complexes

We performed a discovery screen for potential interaction partners of endogenous ZFHX3 using immunoprecipitation (IP) of endogenous ZFHX3 in SH-SY5Y and NSCs, followed by mass spectrometry (MS). This showed enrichment of respectively 83 and 109 proteins compared to an IgG isotype control sample (t-test, FDR=0.05) (Figure 3A, Supplemental Table 5-6). An overlap of 57 ZFHX3 interaction partners was observed in both cell lines (Figure 3B). Among these 57 interactors, SPECC1L is the only known interaction partner of ZFHX3 according to a reference map of human binary protein-protein interactions^36^. This protein is involved in actin-cytoskeleton reorganization and is crucial for proper facial morphogenesis^37^. Mapping the 56 remaining newly identified ZFHX3 binding partners against the STRING database^38^ reveals several ZFHX3-containing protein complexes, such as the mammalian SWI/SNF (mSWI/SNF) complex also known as the BRG1/BRM associated factor (BAF) complex^39^, the cleavage and polyadenylation (CP) complex, the nuclear pore complex (NPC), and the septin complex (Figure 3C). In agreement with the function of these complexes, we show enrichment of the following processes by the gene ontology (GO) enrichment analysis of the ZFHX3 interactome using Metascape^40^: nucleosome assembly, mRNA transport from nucleus to cytoplasm and cytoskeleton-dependent cytokinesis (Supplemental Figure 6). This supports the finding of chromatin remodeling and mRNA binding as enriched biological processes in our transcriptome data mining (see Figure 2E).

**Figure 3:**
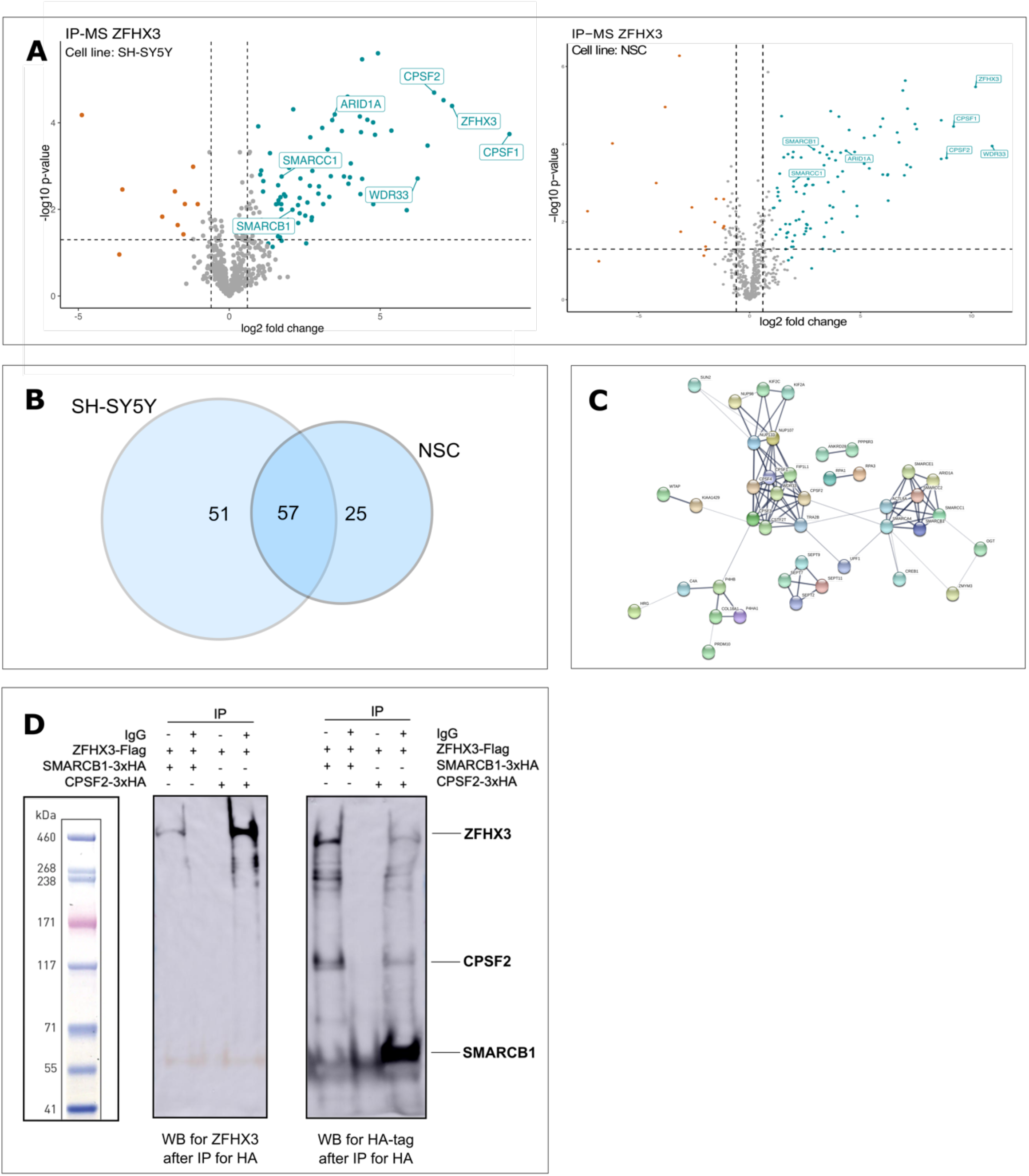
ZFHX3 has a robust protein-protein interaction network of 57 interaction partners. A. Volcano plot showing the enriched ZFHX3 interactor proteins in SH-SY5Y (left: n=83) and NSCs (right: n=109). For each detected protein, the log_2_ fold change (FC) between the ZFHX3 IP-enriched samples and the IgG controls is shown on the X-axis, while the -log_10_ of the p-value is shown on the y-axis. Vertical dashed lines indicated a fold change of abs(0.6). The horizontal dashed line indicates the -log_10_ value of a p-value equal to 0.05. Proteins with an FDR<0.05, log_2_ FC>0.6/<-0.6 and S_0_=1 are respectively highlighted in blue & red. Important members of the BAF and CP complexes are also highlighted, as well as ZFHX3 itself. B. **Overlap of ZFHX3 interaction factors between SH-SY5Y and NSCs IP results**. A total of 57 proteins interacts in both cell lines with ZFHX3. A total of respectively 51 or 25 interaction partners for ZFHX3 were only found in SH-SY5Y or NSCs. **C. ZFHX3 protein-protein interaction network extracted from the STRING 11.0 database.** Shown are only interactors that are connected within a network. Line thickness indicates the strength of data support. The BAF complex, CPSF and CSTF, nuclear pore complex (NPC), and septin complex are novel ZFHX3 containing protein complexes. CPSF and CSTF are two multi-subunit protein complexes responsible for cleavage and polyadenylation^76^. **D. ZFHX3-SMARCB1 and ZFHX3-CPSF2 interaction validated with co-immunoprecipitation in HEK293T cells Left:** Detection of ZFHX3 with anti-ATBF1 antibody after co-immunoprecipitation. **Right:** SMARCB1-3xHA and CPSF2-3xHA were detected with anti-HA antibody on Western Blot after the co-immunoprecipitation. ZFHX3 protein bands were found at a height corresponding with ±460 kDa, SMARCB1-3xHA corresponds to the ±50kDa band and and CPSF2-3xHA to ±91 kDa.

To validate the ZFHX3 protein-protein interactions with the BAF and CP complex, we selected SMARCB1 (subunit of the BAF complex) and CPSF2 (subunit of CP complex), respectively, to perform co-immunoprecipitation, since both were identified in the IP-MS screen as ZFHX3 interactors in SH-SY5Y and NSCs (Figure 3A, Supplemental Table 5&6). In HEK293T, overexpression (Supplemental Figure 7) of a FLAG-tagged ZFHX3 (±404 kDa) with either a 3xHA-tagged SMARCB1 (±50 kDa), or a 3xHA-tagged CPSF2 (±91 kDa), respectively, followed by immunoprecipitation with a HA-tag antibody and Western blot for ZFHX3 corroborates the presence of ZFHX3 among the SMARCB1-3xHA and CPSF-3xHA binding protein extracts (Figure 3D). In addition to the endogenous IP-MS data, these results support ZFHX3 as an interactor of the BAF and CP complexes.

### Loss-of-function variants affecting *ZFHX3* are associated with a methylation profile

Mutations in genes encoding BAF subunits are associated with Coffin-Siris syndrome (CSS), Nicolaides-Baraitser syndrome (NCBRS), Kleefstra syndrome and ASD, commonly referred to as BAFopathies^41^. Aref-Eshgi et al. described specific DNA methylation epi-signatures in blood from patients with BAFopathies^41^. Since ZFHX3 interacts with the BAF complex, we investigated whether *ZFHX3* aberrations associate with a specific methylation profile. To this end, 10 individuals from our patient cohort (six individuals with *ZFHX3* PTVs and four with a (partial) deletion of *ZFHX3*, listed in Supplemental Table 7) were used for probe selection and model construction. We selected 50 control samples matched to the case samples by age, sex, and array type and a set of 208 probes were identified to be differentially methylated between the case and control groups.

The robustness of the selected probes was verified using hierarchical clustering and multidimensional scaling (MDS) models (Figure 4A, B). The set of 208 probes were then used in order to construct Support Vector Machine (SVM) models in order to classify the *ZFHX3* haploinsufficiency cases with more accuracy. Using these models, two Methylation Variant Pathogenicity (MVP) plots were generated, where the first model was constructed by training the 10 case samples against matched controls, and the second model by training the 10 case samples against matched control samples and 75% of other control samples and individuals with other disorders from the EpiSign Knowledge Database (EKD, https://episign.lhsc.on.ca/index.html), to increase the specificity of the model to *ZFHX3* haploinsufficiency. In both models, the remaining samples from the EKD, BAFopathy samples, and three samples with *ZFHX3* missense variants (classified as Variant of Unknown Significance, Supplemental Table 8) were used for testing. The samples with missense variants and the BAFopathy samples received low MVP scores, demonstrating that they did not match the methylation profile identified for *ZFHX3* haploinsufficiency (Figure 4C, D).

**Figure 4.**
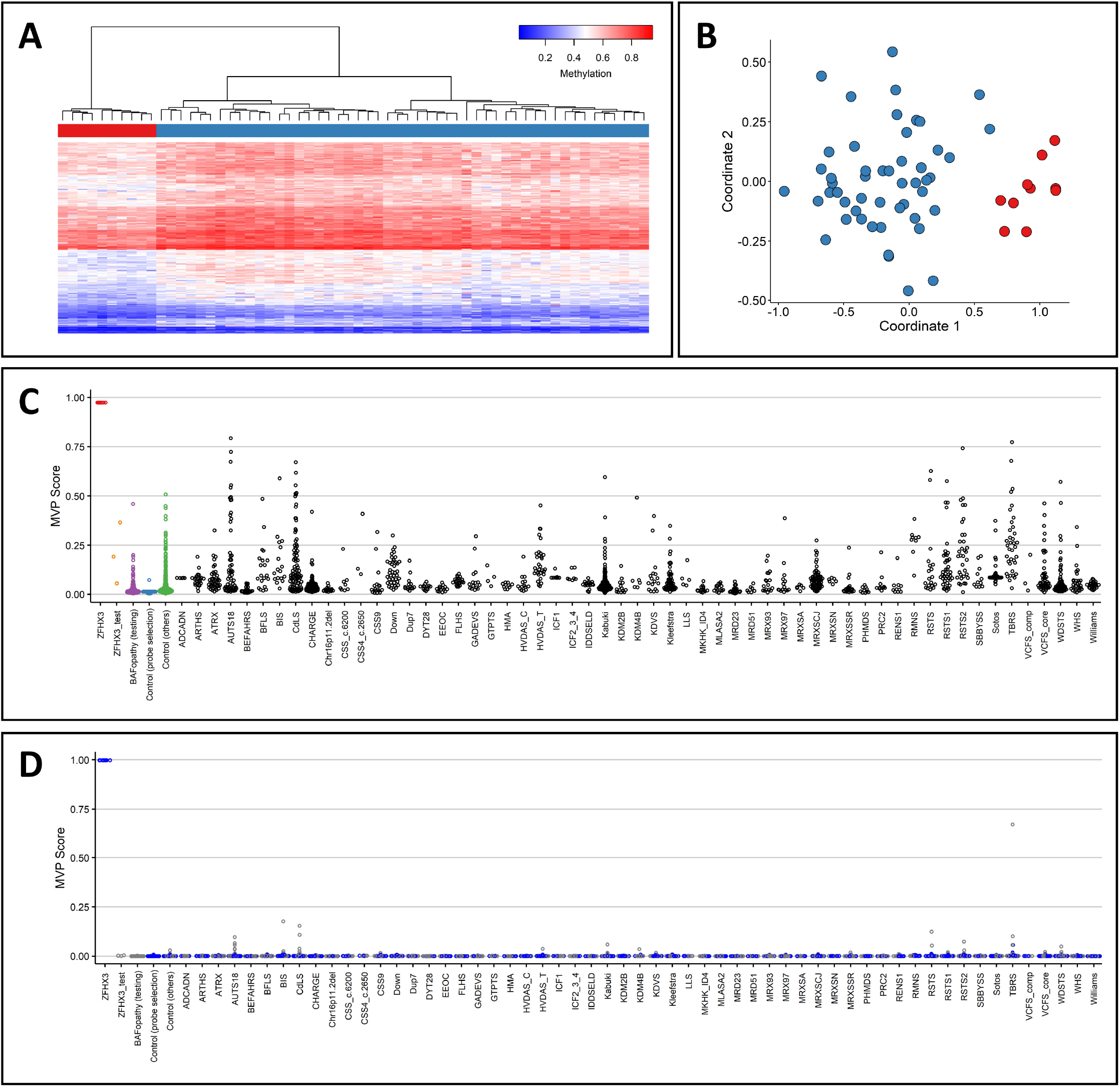
Deletion of *ZFHX3* and *ZFHX3* PTVs are associated with a specific methylation profile. **A.** Hierarchical clustering using the selected CpG sites, where rows represent selected probes and columns indicate samples. Case samples are illustrated with red, and control individuals with blue on the heatmap pane. The heatmap color scale demonstrates methylation levels ranging from blue (no methylation or 0) to red (full methylation or 1). Clear separation between ZFHX3 and control samples is observed. **B.** MDS, demonstrating different methylation pattern between the case and control groups using the selected CpG sites. Red and blue circles represent case and control samples, respectively. **C.** MVP scores generated by the SVM classifier trained using only the 10 ZFHX3 samples and the 50 matched control samples, where the majority of the other control samples and individuals with other disorders received low scores, demonstrating the high specificity of the methylation pattern to ZFHX3 haploinsufficiency. **D.** MVP scores generated by the SVM constructed by training the 10 ZFHX3 samples against matched control samples and 75% of other control samples and samples from other disorders (blue). The remaining 25% of the database samples used for testing (grey) received very low MVP scores, illustrative of the significant improvement in the specificity of the model. The 3 samples with missense variants in *ZFHX3* and the BAFopathy samples were also supplied into the model as testing samples and received low MVP scores, indicating that their methylation pattern is different from that of *ZFHX3* haploinsufficiency.

### ZFHX3 predominantly binds promoters of genes involved in nervous system development

ChIP-seq was performed for endogenous and overexpressed ZFHX3 (see Materials & Methods) in NSCs for which respectively 24,674 and 57,886 peaks were called. Of the 24,674 peaks identified with ChIP-seq for ZFHX3 in NSCs, 22,094 (or 89.5%) were also called in NSCs with ectopic expression of ZFHX3 (Supplemental Table 12). Functional enrichment analysis of these 22,094 overlapping peaks revealed that ZFHX3 predominantly binds to promoter regions (57,18%) (Figure 5B). Furthermore, GO analysis revealed that

ZFHX3 binds to genes involved in axonogenesis, axon development, cell growth and development regulation of the nervous system and neurogenesis (Figure 5C). KEGG pathway enrichment analysis, showed that the proteins encoded by these genes are involved in axon guidance, the Wnt, Hippo and mTOR signaling pathways, all pathways strongly involved the nervous system development^42–44^ (Figure 6D). These results again provide clear indications of a key role for ZFHX3 in neuron and axon development.

**Figure 5.**
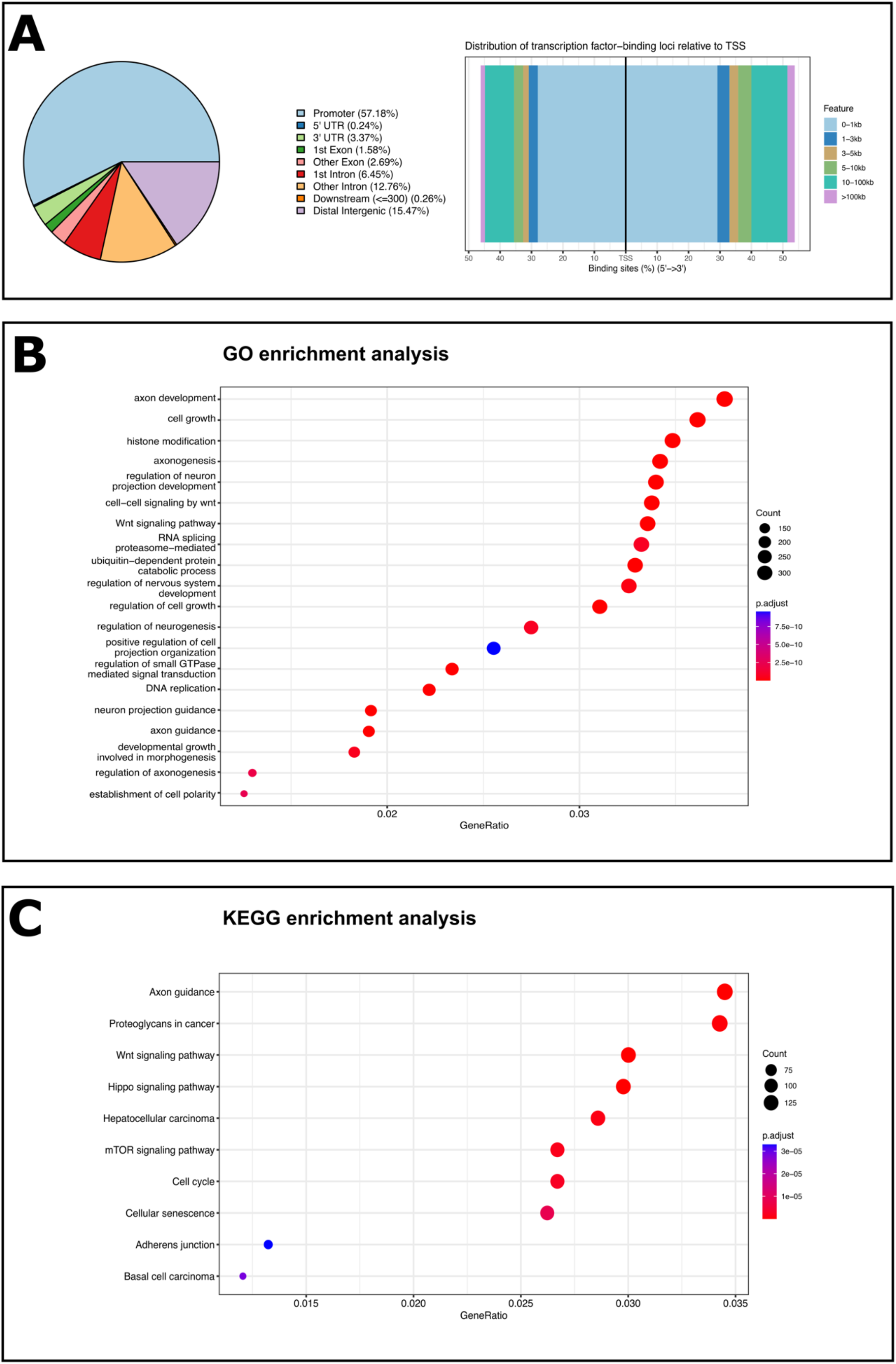
ZFHX3 binds primarily to promoter regions of genes associated with pathways involved in neuron and axon development. B. Left: Annotation of the 22,094 overlapping ChIP-seq peaks. 57,18% of these peaks correspond to promoter regions. The pie chart visualizes the genomic annotation, displaying the percentage of the peaks that reside in/contain the TSS, exonic sequences, 5’UTR, 3’UTR, intronic or intergenic regions. Colour code used to assign each region. **Right: Distribution of the ZFHX3-binding loci relative to** transcription starting sites (**TSS)**. The majority of the peaks falls within 0-1 kb of the TSS, also suggesting that ZFHX3 mainly binds to promoter sequences. Colour code used to represent the distance from TSS in kb. C. **The targets genes of ZFHX3 are involved in axonogenesis, axon development and the regulation of the nervous system, including neurogenesis**. The Gene Ontology (GO) dotplot displays the top 20 enriched biological processes (BP) ranked by gene ratio (# genes related to GO term / total number of significant genes) and the p-adjusted values for these terms (color). The size of the dot represents the gene counts per BP. **D. ZFHX3 plays a key role in neuron and axon developmental pathways.** The Kyoto Encyclopedia of Genes and Genomes (KEGG) dotplot provides all enriched biological pathways ranked by gene ratio (# genes related to GO term / total number of significant genes) and the p-adjusted values for these terms (color). The size of the dot represents the gene counts per pathway.

### zfh2, the ZFHX3 orthologue, is a key gene for neurodevelopment in flies

The suggested ortholog of *ZFHX3* in *Drosophila melanogaster* is *zfh2* (FlyBase ID: <u>FBgn0004607</u>). The gene is well conserved with a DIOPT score of 12/16 (DIOPT v.8.0, Figure 6A). The protein encoded by *zfh2* shows 23% identity and 33% similarity to the human ZFHX3 across the entire protein (Figure 6A&B). To experimentally prove functional conservation of both proteins in flies, we generated fly lines overexpressing both zfh2 (UAS-zfh2) and ZFHX3 (UAS-ZFHX3). We opted to use the PhiC31 integrase-mediated insertion, minimizing the influence of genomic background on expression levels. Ectopic expression of both zfh2 or ZFHX3 in wing discs during development, using apterous GAL4, caused defective midline closure resulting in cleft thorax (Figure 6C). Overexpression of both proteins thus phenocopies each other, pointing towards functional conservation. Next, we investigated whether zfh2 is expressed in developing neurons. We therefore performed immunohistochemistry on third instar larval brains and observed that zfh2 is present in the nucleus of neurons in control (W1118) larval brains (Supplemental Figure 9). This was confirmed using a zfh2-CRIMIC line, where the endogenous zfh2 gene is replaced by a GAL4 reporter cassette, revealing promoter activity upon crossing it to a UAS-RFP reporter line. Finally, also in a BAC line with overexpression of a GFP-tagged zfh2 under control of its own regulatory elements we observed a similar expression pattern (Figure 6D). From this latter line, chromatin immunoprecipitation (ChIP) sequencing data using whole embryos are publicly available^45^. Functional enrichment analysis revealed that zfh2 binds to genes involved in (neural) development and differentiation (Figure 6E) pointing again towards a key function of zfh2 in neurons. Moreover, general knockdown (Actin-Gal4), using three independent RNAi lines as well as neuron-specific knockdown (ELAV-Gal4 and Nsyb-Gal4) or zfh2 overexpression, all lead to developmental lethality (Figure 6E, Supplemental Table 10). Correct dosing of zfh2 during development and more specifically balanced zfh2 levels in neurons thus seems to be of crucial importance for normal development (Figure 6F, Supplemental Table 10).

**Figure 6.**
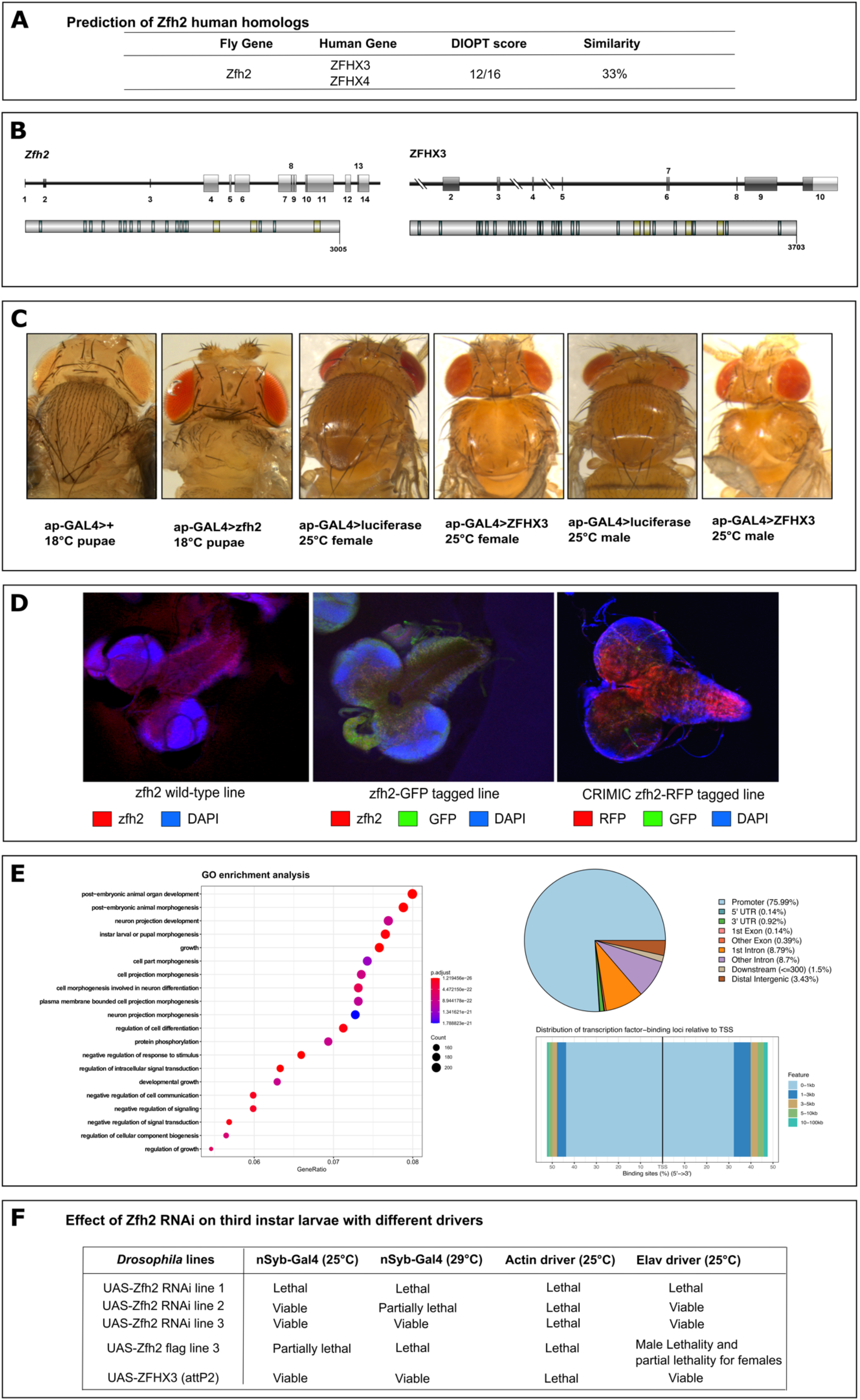
zfh2 is the Drosophila orthologue of ZFHX3. **A.** *zfh2*, *ZFHX3*’s ortholog in flies, has a high homology score of 12/16 (DIOPT v8.0^77^). **B. Gene and protein structure of fly *zfh2*.** Left: The coding exons of *zfh2* are shown as dark grey boxes, and untranslated regions (UTR) in light grey. Below the zfh2 protein is shown with its specific domains. Right: The coding exons of *ZFHX3* are shown as dark grey boxes, and untranslated regions (UTR) in light grey. Below the zfh2 protein is shown with its specific domains. **C. Ectopic expression of zfh2 or ZFHX3 in wing discs results in defective midline closure resulting in cleft thorax.** From left to right: 1) a control luciferase line with the apterous driver at 18°C in the pupae stage, 2) zfh2 ectopic expression with the apterous driver at 18°C in the pupae stage, 3) control luciferase female fly with the apterous driver at 25°C, 4) ZFHX3 ectopic expression in female fly with the apterous driver at 25°C, 5) control luciferase male fly with the apterous driver at 25°C and 6) ZFHX3 ectopic expression in male fly with the apterous driver at 25°C. All pupae and flies with ectopic expression of zfh2 or ZFHX3 show clear defective midline closure. **D. Expression of zfh2 (RFP) in the third instar larval brain in three different fly lines (zfh2 wild-type, zfh2-GFP tagged and CRIMIC zfh2-RFP tagged lines)**. GFP is shown in green, zfh2:RFP is shown in red and DAPI is shown in dark blue. All images show that zfh2 localizes to the nucleus of the neurons. **E. Functional enrichment analysis of zfh2 ChIP-seq peaks in whole *Drosophila melanogaster* embryos**. **Left**: The Gene Ontology (GO) dotplot displays the top 20 enriched biological processes (BP) ranked by gene ratio (# genes related to GO term / total number of significant genes) and the p-adjusted values for these terms (color). The size of the dot represents the gene counts per BP. The proteins encoded by these genes seem to play a role in several developmental processes. **Right Top**: **Annotation of the 4,344 ChIP-seq peaks.** 75,44 % of these peaks correspond to promoter regions. The pie chart visualizes the genomic annotation, displaying the percentage of the peaks that reside in the TSS, exonic regions, 5’UTR, 3’UTR, intronic or intergenic regions. Colour code used for each region. **Right Bottom: Distribution of the zfh2-binding loci relative to TSS**. The majority of the peaks falls within 0-1 kb of the TSS, also suggesting that zfh2 mainly binds to promoter sequences. Colour code used to represent the distance from TSS in kb.. **F. Effect of zfh2 RNAi in third instar larvae with different drivers.** All RNAi lines were lethal after an overall knockdown of zfh2 with an Actin driver at 25°C. Using neuronal drivers nSyb and elav at 25°C, lethality was observed in the TRIP zfh2 RNAi line.

## Discussion

We identified loss of function variation in *ZFHX3* as the cause for a novel NDD, characterized by developmental delay and/or intellectual impairment and/or behavioral problems, feeding difficulties, postnatal growth retardation, and recognizable facial features. Patients with larger microdeletions containing *ZFHX3* and other coding genes present with a similar phenotype, albeit with more pronounced developmental delay, pointing towards a primary role for ZFHX3 in these microdeletions.

Proband 1 harbors a multigenic deletion also including *AP1G1* that encodes the AP1γ1 protein, important for the formation of clathrin coated vesicles. De *novo* PTVs and missense variants as well as bi-allelic missense variants affecting *AP1G1* have recently been associated with the Usmani-Riazuddin syndrome (OMIM #619467)^46^. As the consequences on AP1γ1 protein levels, protein aggregation and clathrin formation are divergent for the different variants, it remains unclear if the disease mechanisms result from pure AP1G1 loss-of-function. Moreover, since the phenotype associated with *de novo AP1G1* mutations seems relatively similar to the phenotype observed for *ZFHX3* LoF, a potential effect of the *AP1G1* deletion cannot be excluded in proband 1.

All identified single nucleotide and indel variants are truncating and likely activate the NMD pathway, as they are situated more than 50-55 nucleotides upstream of the final exon-exon junction. Probands with a *ZFHX3* microdeletion are clinically indistinguishable from probands with *ZFHX3* truncating variants. Moreover, preliminary data provided promising evidence for the existence of a shared methylation profile for individuals harboring either a microdeletion including ZFHX3 or a PTV in ZFHX3, suggesting haploinsufficiency of *ZFHX3* as the underlying mechanism in both. In addition to these LoF variants, we tested three missense variants’ samples for the methylation profile. They however did not reveal a similar methylation profile, suggesting the molecular mechanisms of the different protein variants are not similar. Of note, in one of the individuals with a missense variant, a structural variant disrupting SON was later identified via optical mapping, matching the phenotype. In previous studies, (compound) heterozygous missense variants of *ZFHX3* were reported in probands with epilepsy^20, 47^ and behavioral issues, but normal IQ^48, 49^. Consequently, we cannot rule out that missense variation in *ZFHX3* could associate with epilepsy, rather than with a primary cognitive phenotype, despite the observation that missense variation in ZFHX3 is not restrained (z-score -1.23 in gnomAD (release v2.1.1). Recently Zhang et al. reported two *de novo ZFHX3* missense variants respectively in an individual with non-syndromic esophageal atresia (p.(Pro534Arg)) with or without tracheoesophageal fistula (EA/TEF) and a patient with EA/TEF as part of his VATER/VACTERL association (p.(Ala2126Val). They suggest *ZFHX3* as a putative candidate gene for EA/TEF since it has a high expression in mouse embryonic foregut tissue and in the pharyngeal arches in zebrafish larvae. EA/TEF was however not observed in our patient cohort.

Further work on the different types of variants in *ZFHX3*, including phenotypic delineation, is ongoing to enable development of a sensitive and specific diagnostic biomarker, that would be beneficial for the interpretation of *ZFHX3* aberrations.

Functional enrichment analysis of ZFHX3 and zfh2 ChIP-seq data from respectively human NSCs and whole embryos *Drosophila* shows that both proteins predominantly bind to promoter regions. The functional enrichment analysis in both ChIP-sequencing data sets indicates functional conservation of the homologs as both bind genes involved in neural and neural crest development and migration, including genes encoding key members of the Hippo/YAP pathway^44^. A member of the Hippo signalling pathway, YAP, was found to play crucial role in regulating neural progenitor cell number by influencing proliferation, fate choice, and cell survival^50^. In addition, the Hippo pathway seems to control the proliferative potential of neuroblasts and overall brain size in the *Drosophila* brain^51^. Other important pathways and processes influenced by ZFHX3 functioning are Wnt/β-catenin and mTOR signaling and cell cycling. Wnt/β-catenin signaling is crucial during neurodevelopment and function of the CNS, including neurogenesis, cell pluripotency and fate decisions^42, 52–55^, while dysregulated mTOR signaling has been associated with impaired neurogenesis, neurodevelopmental and neuropsychiatric disorders^43, 56, 57^. In addition, ZFHX3 has already been shown to modulate cell cycling, triggering cell cycle arrest in neuronal progenitor cells to induce neuronal differentiation^58^.

The *Drosophila* orthologue zfh2 is expressed in neurons and we observed that general knockdown using three zfh2 RNAi lines and neuronal knockdown with one RNAi line and the UAS-zfh2 line, results in developmental lethality, confirming a key role for zfh2 in development. Similarly, mice with a heterozygous truncating deletion of exon 7 and 8 in *Zfhx3* show growth retardation and preweaning mortality^59^. In addition, defects in other members of the ZFHX gene family have been associated with neuropsychiatric or neurodevelopmental phenotypes in humans and mice. In humans, the paralog *ZFHX4* is likely the major contributing gene in the 8q21.11 microdeletion syndrome, associated with intellectual disability^2, 60^. *Zfhx2*-deficient mice show behavioral abnormalities (hyperactivity, depression, and anxiety-related behavior), but normal cognition^41^.

In agreement with the ChIP-seq data, the spatiotemporal expression of *ZFHX3* in the developing mouse^61^, rat^58^, fruit fly^62^ and human brain, as well as in differentiating neuronal progenitor cells supports a role for neuronal differentiation^13, 63^. Within the *Drosophila* neuroblasts, the temporal identity of zfh2 is well described as a later born neuron marker^62^. In our study, we confirm the expression of zfh2 protein in neurons of third instar larval brains. In the single-cell data from recently published Fly Cell Atlas^64^, zfh2 was mainly localized in neurons, sensory neurons and glial cells in fly adult brains. The temporal patterning is similar to that of Zfhx3 within mice striatum from previously published single-cell sequencing data^16^.

The complex structure of ZFHX3, containing 4 homeodomains, suggests dynamic functions in diverse biological processes^5^. In line with this, we observed several major interaction partners. Its interaction with several subunits of the BAF complex suggests a role in chromatin remodeling. The BAF complex is a chromatin remodeling complex that consists of up to 15 subunits. It mediates DNA accessibility by repositioning of nucleosomes in an ATP-dependent manner, hence regulating gene expression. Not surprisingly, pathogenic variants in several subunits of the BAF complex were found in NDDs within the spectrum of Coffin-Siris syndrome and Nicolaides-Baraitser syndrome^65, 66^ commonly coined BAFopathies. Of note, ectopic expression in the wing disc of Osa, a confirmed component of the Brahma complex, the *Drosophila* homolog of the BAF complex, results in alterations of the bristle formation and defects in the midline of the notum, scutellum and abdomen; similarly to our observations in flies with ZFHX3 ectopic expression^67^. This phenotype has been implicated with the dysregulation of the JNK signaling pathway^68^. However, to unravel the role of zfh2 and Osa in this pathway, further investigation is needed.

Messenger RNA transcripts undergo several processing events including addition of a 5’-cap, splicing, and 3’-end cleavage and polyadenylation before transportation to the cytoplasm^69^. In addition to their role in transcription regulation, chromatin remodelers may influence pre-mRNA splicing and 3’-end processing^70^. Previous studies showed a physical and functional interaction between the BAF complex and cleavage and polyadenylation factors^71^. Furthermore, many 3’ end processing factors (CPSF, CSTF and PABP), nucleoporins and a subset of the SWI/SNF chromatin remodeling complex are identified in supraspliceosomes (or polyspliceosomes) in HeLa and chicken cells^72^. Since we identify ZFHX3 as an interactor with these 3’ end processing factors, this raises the intriguing hypothesis that ZFHX3 facilitates interconnection of these mRNA processing activities.

Furthermore, we identify an interaction between ZFHX3 and the septin complex. The cytosolic septin complex serves an important role in various cellular processes, pertinent for several neuronal functions, such as axon dynamics and growth, and dendrite formation^73^. This is in line with the cytoskeletal function of the previously identified interaction partner SPECC1L^36^. In previous studies, decreased SPECC1L expression has been associated with cytoskeletal and cell migration malfunctioning^74^. Heterozygous pathogenic SPECC1L variants are associated with impaired facial morphogenesis resulting in facial clefts and prominent hypertelorism^37, 75^. The role of SPECC1L as a critical organizer of vertebrate facial morphogenesis also confirmed in other species such as zebrafish and *Drosophila melanogaster*^37^. Of note, knockdown of CG133GG, the Drosophila orthologue of SPECCL1, in the wing imaginal disc results in a high percentage of uninflated, crumpled wings and midline thorax closure defects, indicative of cell migration and adhesion defects^37^.

In conclusion, we report loss-of-function variants in *ZFHX3* as a cause for syndromic ID, associated with a specific DNA methylation profile. ZFHX3 is essential for neuronal differentiation and brain development due to its functions in chromatin remodelling and mRNA processing.

## Data Availability

Public data was used from:
https://apps.kaessmannlab.org/evodevoapp/
https://www.gtexportal.org/home/
http://www.brainspan.org
https://bioinf.eva.mpg.de/shiny/sample-apps/scApeX/
https://shcheglovitov.shinyapps.io/u_brain_browser/
https://gnomad.broadinstitute.org/about
https://doi.org/doi:10.17989/ENCSR259WOE
The analyzed IP-MS data and the overlapping peaks between endogenous and ectopic ZFHX3 ChIP-seq is available in the Supplementary Material.
Some of the DNA methylation datasets used in this study are publicly available and may be obtained from gene expression omnibus (GEO) using the following accession numbers. GEO: GSE116992, GSE66552, GSE74432, GSE97362, GSE116300, GSE95040, GSE 104451, GSE125367, GSE55491, GSE108423, GSE116300, GSE 89353, GSE52588, GSE42861, GSE85210, GSE87571, GSE87648, GSE99863, and GSE35069. These include DNA methylation data from patients with Kabuki syndrome, Sotos syndrome, CHARGE syndrome, immunodeficiency-centromeric instability-facial anomalies (ICF) syndrome, Williams-Beuren syndrome, Chr7q11.23 duplication syndrome, BAFopathies, Down syndrome, a large cohort of unresolved subjects with developmental delays and congenital abnormalities, and also several large cohorts of DNA methylation data from the general population. Remaining methylation data is not publicly available due to institutional and ethics restrictions.

https://www.gtexportal.org/home/

http://www.brainspan.org

https://bioinf.eva.mpg.de/shiny/sample-apps/scApeX/

https://shcheglovitov.shinyapps.io/u_brain_browser/

https://gnomad.broadinstitute.org/about

https://doi.org/doi:10.17989/ENCSR259WOE

## Acknowledgements

First of all, we would like to thank the reported families for their cooperation in this study. We are grateful for the group of Peter Ponsaerts, University of Antwerp, by providing us iPSC-derived NSCs in order to start the culture in our own at our lab. We would also like to thank the research group of Prof Patrick Nolan (MRC Harwell, Harwell Science and Innovation Campus, Oxfordshire, United Kingdom) for providing us their ZFHX3 custom antibody and Prof. Chris Doe (Institute of Neuroscience, Institute of Molecular Biology, Howard Hughes Medical Institute, University of Oregon, USA) for the zfh2 custom antibody. Concerning the *Drosophila* work, we would like to thank Prof. Fernando J Díaz-Benjumea (Centro de Biología Molecular Severo Ochoa, Universidad Autónoma de Madrid (UAM), Spain) for kindly providing their *Drosophila* Zfh2 RNAi line.

## Supplementary material

Supplementary material is available in the final published manuscript.

## Appendix 1

### ZFHX3 consortium

Pankaj Agrawal (Boston Children’s Hospital, MA 02115, USA), Daryl Armstrong Scott (Baylor College of Medicine, Texas Children’s Hospital, TX 77030, USA), Elizabeth Barkoudah (Boston Children’s Hospital, Department of Neurology, MA 02115, United States), Melissa Bellini (USSD Laboratorio Genetica Medica, ASST Papa Giovanni XXIII, 24127 Bergamo BG, Italy), Claire Beneteau (CHU Nantes, Service de Génétique Médicale, 44000 Nantes, France), Kathrine Bjørgo (Department of Medical Genetics, Oslo University Hospital, 0424 Oslo, Norway), Alice Brooks (Clinical Genetics, Erasmus Medical Center, 3000 CA Rotterdam, The Netherlands), Natasha Brown (Victorian Clinical Genetics Services, Murdoch Children’s Research Institute, Melbourne, 3052, Australia), Alison Castle (PGY5 Medical Genetics and Genomics, CHEO, ON K1H 8L1, Canada), Diana Castro (UT Southwestern Medical Center, Dallas, Texas 75390 USA), Odelia Chorin (Institute of Rare Diseases, Edmond & Lily Safra Hospital for Children, Tel Hashomer, 52621 Ramat Gan, Israel), Mark Cleghorn (Victorian Clinical Genetics Services, Murdoch Children’s Research Institute, Melbourne, 3052, Australia), Emma Clement (Clinical Genetics Department, Great Ormond Street Hospital, London WC1N 3JH, United Kingdom), David Coman (Queensland Children’s Hospital, South Brisbane QLD 4101, Australia), Carrie Costin (Akron Children’s Hospital, OH 44308, USA), Koen Devriendt (Centre for Human Genetics, KU Leuven, Leuven, 3000, Belgium), Dexin Dong (Peking Union Medical College Hospital, Chinese Academy of Medical Sciences, Beijing, Dongcheng, China 100006), Annika Dries (Stanford University, 94305, USA), Tina Duelund Hjortshøj (Kennedy Center, Department of Clinical Genetics, Copenhagen University Hospital, Rigshospitalet, DK-2600 Glostrup, Denmark), David Dyment (PGY5 Medical Genetics and Genomics, CHEO, ON K1H 8L1, Canada), Christine Eng (Baylor College of Medicine, TX 77030, USA), Casie Genetti (Boston Children’s Hospital, MA 02115, USA), Siera Grano (Texas Scottish Rite Hospital for Children, Dallas, TX 75219 USA), Peter Henneman (Department of Human Genetics, Amsterdam Reproduction & Development Research Institute, Amsterdam University Medical Centers, Amsterdam, 1105 AZ, The Netherlands), Delphine Heron (Hôpitaux Universitaires Pitié Salpêtrière - Charles Foix, 75013 Paris, France), Katrin Hoffmann (Division of Genetics, Children’s Hospital, University of Freiburg, 79106 Freiburg, Germany; Institute of Human Genetics, University Hospital Halle, Martin Luther University Halle-Wittenberg, 06112 Halle, Germany), Jason Hom (Stanford University, 94305, USA), Haowei Du (Baylor College of Medicine, TX 77030, USA), Maria Iascone (USSD Laboratorio Genetica Medica, ASST Papa Giovanni XXIII, Bergamo, Italy), Bertrand Isidor (CHU Nantes, Service de Génétique Médicale, Nantes, France), Irma E. Järvelä (Department of Medical Genetics, University of Helsinki, 00014, Finland), Julie Jones (Greenwood Genetic Center, North Charleston, SC, 29418 USA), Boris Keren (Hôpitaux Universitaires Pitié Salpêtrière - Charles Foix, 75013 Paris, France), Mary Kay Koenig (UTHealth Houston, 77030, United States), Jürgen Kohlhase (Division of Genetics, Children’s Hospital, University of Freiburg, 79106 Freiburg, Germany), Seema Lalani (Baylor College of Medicine, Texas Children’s Hospital, TX 77030, USA), Cedric Le Caignec (CHU Nantes, Service de Génétique Médicale, 44000 Nantes, France), Andi Lewis (Baylor College of Medicine, Texas Children’s Hospital, TX 77030, USA), Pengfei Liu (Baylor College of Medicine, TX 77030, USA), Alysia Lovgren (Center for Mendelian Genomics, Broad Institute, Cambridge, MA 02142, USA), James R. Lupski (Baylor College of Medicine, Texas Children’s Hospital, TX 77030, USA), Mike Lyons (Greenwood Genetic Center, North Charleston, SC, 29418 USA), Philippe Lysy (Pediatric Endocrinology Unit, Cliniques Universitaires Saint Luc, Pôle PEDI, 1200 Woluwe-Saint-Lambert, Belgium; Institut de Recherche Expérimentale et Clinique, Université Catholique de Louvain, 1200 Woluwé-Saint-Lambert, Belgium), Melanie Manning (Stanford University, 94305, USA), Carlo Marcelis (Department of Human Genetics, Radboudumc, Nijmegen, 6525 GA, The Netherlands), Scott Douglas McLean (Baylor College of Medicine, Children’s Hospital of San Antonio, 78207 USA), Sandra Mercie (CHU Nantes, Service de Génétique Médicale, 44000 Nantes, France), Mareike Mertens (University of Halle (Saale) Medical Care Center, Halle (Saale), Germany and Institute of Human Genetics, University of Leipzig Medical Center, Leipzig, Germany), Arnaud Molin (CHU Caen, CS 30001 - 14 033 Caen Cedex 9, Normandie, France), Mathilde Nizon (CHU Nantes, Service de Génétique Médicale, 44000 Nantes, France), Kimberly Margaret Nugent (Baylor College of Medicine, Children’s Hospital of San Antonio, 78207 USA), Susanna Öhman (Kårkullla Joint municipal authority, 21610 Kirjala, Finland), Melanie O’Leary (Center for Mendelian Genomics, Broad Institute, Cambridge, MA 02142 USA), Rebecca Okashah Littlejohn (Baylor College of Medicine, Children’s Hospital of San Antonio, 78207 USA), Florence Petit (Pôle de Biologie Pathologie Génétique, CHU de Lille - Centre de Biologie Pathologie Génétique, 59037, France), Rolph Pfundt (Department of Human Genetics, Radboud University Medical Center, Nijmegen, 6525 GA, The Netherlands), Lorraine Pottocki (Baylor College of Medicine, Texas Children’s Hospital, TX 77030, USA), Annick Raas-Rotschild (Institute of Rare Diseases, Edmond & Lily Safra Hospital for Children, Tel Hashomer, 52621 Ramat Gan, Israel), Kara Ranguin (CHU Caen, CS 30001 - 14 033 Caen Cedex 9, Normandie, France), Nicole Revencu (Center for human genetics, Cliniques universitaires Saint Luc and Université catholique de Louvain, Brussels, Belgium), Jill Rosenfeld (Baylor College of Medicine, TX 77030, USA), Lindsay Rhodes (GeneDx, MD 20877 USA), Fernando Santos Simmaro (INGEMM, Instituto de Genética Médica y Molecular, IdiPAZ, Hospital Universitario la Paz, Universidad Autónoma de Madrid (UAM), 28029 Madrid, Spain), Karen Sals (Exeter Genomics Laboratory, Royal Devon and Exeter NHS Foundation Trust, Exeter EX2 5DW, UK) Jolanda Schieving (Amalia Children’s Hospital, Radboud University Medical Center, Nijmejen 6525 GA, The Netherlands), Isabelle Schrauwen (Columbia University Irving Medical Center, NY 10032, United States), Janneke H.M. Schuurs-Hoeijmakers (Radboud University Medical Center, Nijmegen 6525 GA, The Netherlands), Eleanor G. Seaby (Faculty Of Medicine, University of Southampton, Southampton, SO16 6YD, UK), Ruth Sheffer (Department of Genetics and Metabolic Diseases, Hadassah-Hebrew University Medical Center, 91120 Jerusalem, Israel), Lot Snijders Blok (Radboud University Medical Center, 6525 GA, The Netherlands), Kristina Sorensens (Department of Clinical Genetics, Odense University Hospital, 5000 Odense C, Denmark), Siddharth Srivastava (Boston Children’s Hospital, Department of Neurology, MA 02115, United States), Zornitza Stark (Victorian Clinical Genetics Services, Murdoch Children’s Research Institute, Melbourne, 3052, Australia), Radka Stoeva (CHU Nantes, Service de Génétique Médicale, 44000 Nantes, France), Chloe Stutterd (Victorian Clinical Genetics Services, Murdoch Children’s Research Institute, Melbourne, 3052, Australia), Pernille Mathiesen Torring (Department of Clinical Genetics, Odense University Hospital, 5000 Odense C, Denmark), Olivier Vanakker (Center for Medical Genetics Ghent, Ghent University Hospital, 9000, Belgium), Liselot van der Laan (Department of Human Genetics, Amsterdam Reproduction & Development Research Institute, Amsterdam University Medical Centers, Amsterdam, 1105 AZ, The Netherlands), Athina Ververi (Clinical Genetics Department, Great Ormond Street Hospital, London WC1N 3JH, United Kingdom), Pablo Villavicencio-Lorini (Institute of Human Genetics, University Hospital Halle, Martin Luther University Halle-Wittenberg, 06112 Halle, Germany; MVZ UKH gGmbH, Center for Pediatrics and Human Genetics, 06120 Halle, Germany), Marie Vincent (CHU Nantes, Service de Génétique Médicale, 44000 Nantes, France), Dorothea Wand (Department Medical Genetic and Pathology, University Hospital of Basel (USB), CH-4031 Basel, Switzerland), Marja Wessels (Clinical Genetics, Erasmus Medical Center, 3000 CA Rotterdam, The Netherlands), Sue White ((Victorian Clinical Genetics Services, Murdoch Children’s Research Institute, Melbourne, 3052, Australia), Monica H Wojcik (Boston Children’s Hospital, MA 02115, United States), Nan Wu (Peking Union Medical College Hospital, Peking Union Medical College and Chinese Academy of Medical Sciences, Beijing, Dongcheng, China 100006), (Peking Union Medical College Hospital, Peking Union Medical College and Chinese Academy of Medical Sciences, Beijing, Dongcheng, China 100006), Sen Zhao (Peking Union Medical College Hospital, Chinese Academy of Medical Sciences, Beijing, Dongcheng, China 100006).

