## Supplemental Methods and Figures for "A novel neurodevelopmental syndrome caused by loss-of-function of the Zinc Finger Homeobox 3 (ZFHX3) gene"

### Supplementary Material

### Supplemental Methods

##### Cell culture

The naïve hESCs were cultured on CF-1 irradiated Mouse Embryonic Fibroblasts (MEFs) 2M (A34180, Gibco) in conditioned GDN medium consisting of KO-DMEM (10829018, Invitrogen) containing 20% knockout serum replacement (10828028, Invitrogen), 1% non-essential amino acids (11140050, Invitrogen), 1% Penicillin/Streptomycin (15140122, Invitrogen), 0.1 mM L-glutamine (25030024, Invitrogen) and 0.1mM β-mercaptoethanol (31350010, Invitrogen). The medium is supplemented with 12 ng/ml bFGF (100–18B, Peprotech), 1000U recombinant human LIF (L5283, Sigma) and the small molecules 1µM PD0325903 (13034, Sanbio), 3µM CHIR99021 (1386, Axon Medchem), 10µM Forskolin (F6886, Sigma) and 50 ng/ml ascorbic acid (A8960, Sigma). The naïve colonies were passaged as single cells using 0.05% trypsin/EDTA (25300054, Invitrogen) every three days and were re-plated on inactivated MEFs. To convert the hESCs on feeder cells to feeder-free conditions, the naïve hESCs were passaged with 0.05% trypsin/EDTA (25300054, Invitrogen) onto Matrigel coated plates with conditioned GDN medium (protocol described above) and cultured at hypoxic conditions (37°C, 5% CO2 and 5% 02). On day 1, half of the GDN medium was replaced by the RSeT$^{TM}$ Feeder-Free medium (STEMCELL technologies). Every second day, a full medium change was performed with RSeT$^{TM}$ Feeder-Free medium. On day seven, colonies were passaged with Accutase (Thermofischer Scientific, A1110501) onto freshly made Matrigel coated plates. Every other day, fresh RSeT$^{TM}$ feeder-free medium was added and cells were passaged every third or fourth day (depending on the cell density).

##### Immunofluorescence

Phosphate buffered saline (PBS) was used for all washing steps on all cell types. Autofluorescence quenching of the cells was performed with 50mM NH4Cl for 10 min, followed by permeabilization with 0.2% triton X-100, for 10 min. Non-specific antibody binding was avoided by means of a blocking buffer containing 3% goat serum (GS) (Agilent cat. no. X0907), as well as 1% bovine serum albumin (BSA) in a phosphate buffered saline, and 0.1% Tween 20 (PBS-T) solution, for 30 min at room temperature. The primary antibody, rabbit anti-ZFHX3 (HPA059353, Sigma-Aldrich, 1/100), was added to the cells and incubated overnight at 4°C. The fluorescently labelled secondary antibody, 2 µg of goat anti-rabbit IgG AlexaFluor 488 (A-11034, ThermoFisher) diluted in a Hoechst (brand) solution with blocking buffer, was then added for 1h at room temperature, whilst avoiding light exposure. Cover slips were mounted onto glass slides, and propyl gallate mounting media (SigmaAldrich) were added onto the cover slip.

**Immunoprecipitation followed by mass-spectrometry (IP-MS)**

Protein from $2x10^7$ cells were extracted as mentioned above (Materials). Next, 2µg antibody (ZFHX3 antibody (D1-120, MBL Life Science), Anti-Human IgG antibody (ab2410, Abcam)) was added and after four hours, 20 µl extensively washed protein A Ultralink resin beads (Thermo Scientific) were added, further rotated overnight. The samples were centrifuged and washed five times with 50mM digestion buffer (50mM TrisHCl pH8, 2mM CaCl2) to remove unbound proteins. Beads containing antibody and proteins of interest were resuspended in 175 µl digestion buffer. For IP, 25µl of samples were incubated at 95°C for 10 min with 40 µl of 2x laemmli buffer at 1000 rpm to elute the proteins. Supernatants containing the proteins of interest were used for western blot analysis. For IP-MS, the remaining 150 µl of the samples was incubated for 4 hours with 1 µg trypsin (Promega) at 37℃, while shaking. Beads were removed by centrifugation at maximum speed and proteins were digested again with 1 µg of trypsin, overnight at 37℃. The peptide mixture was acidified with 1% TFA and purified on Omix C18 tips (Agilent). Next, the samples were dried and re-dissolved in 20 µl 0.1% formic acid in water/acetonitrile (98:2, v/v) of which 2 µl was injected for LC-MS/MS analysis on an Ultimate 3000 RSLC nano LC (Thermo Fisher Scientific, Bremen, Germany) in-line connected to a Q Exactive mass spectrometer (Thermo Fisher Scientific). Data analysis was performed with MaxQuant algorithm (version 1.6.3.4) with default search settings including a false discovery rate set at 1% on both the peptide and protein level. Spectra were searched against the human proteins in the Swiss-Prot Reference Proteome database (database release version of January 2019) supplemented with the sequence of recombinant protein A from *Staphylococcus aureus*.

To compare protein intensities between the IgG and ZFHX3 samples, a t-test was performed (FDR=0.05 and $S_0=1$). Quantified proteins and the results of the t-tests are listed in Supplemental Tables 5 (SH-SY5Y) & 6 (NSCs). The fold change (in $\log_2$) of each protein between the 3 IgG and 3 ZFHX3 samples per cell line is shown in column C, while the values in column B indicate the statistical significance (-log P value).

**Expression Constructs, Transfection, Cell harvesting, RNA & protein extraction**

The ZFHX3-FLAG plasmid was ordered from AddGene (pcDNA3.1-FLAG-ATFB1, (#40927). N-term 3xHA-tag followed the ORF in the new designed SMARCB1-3xHA or CPSF2-3xHA expression vector were designed via Gateway Cloning (Gateway Technology, Gateway LR reaction, Invitrogen).

Transfection complexes were prepared according to the Lipofectamine 3000 manufacturer's instructions. Appropriate amounts of the above-described plasmid DNA and P3000 buffer (Lipofectamine 3000 protocol), were diluted and added to the Lipofectamine 3000 solution (Thermofisher Scientific) in a 1:1 ratio, depending on the amount of desired transfected cells. At room temperature and within a 15-minute incubation period, the complex was added to the corresponding amount of HEK293T cells, which were seeded for 24 hours before the transfection. Cells were incubated upon normoxic conditions at 37°C.

**Co-immunoprecipitation (co-IP)**

Immunoprecipitation was performed according to the Dynabeads™ Protein G (Invitrogen) manufacturer's instructions. 300µl lysate was added to the antibody beads complex and incubated overnight at 4°C. Protein complexes were eluted with 20 µl glycine buffer (pH 2.8) and determined using Western Blot. As a negative control for the co-immunoprecipitation (co-IP), 300µl per cell lysate was added to Dynabeads™ protein G, coupled to non-specific IgG antibodies.

The anti-HA.11 Epitope Tag antibody (1/1000, Clone 16B12, Biolegend) was used to enrich the capture of the SMARCB1-HA tagged overexpressed protein and the CPSF2-HA tagged overexpressed protein within the co-IP.

**Western Blot**

Western blot was performed according to an in-house protocol. Either protein lysates or eluted proteins after co-IP were subjected to NuPage gel electrophoresis, subsequently transferred to a nitrocellulose membrane, and detected using the appropriate antibodies: anti-ZFHX3 (custom antibody (Parsons et al., 2015)) kindly provided by Prof. Patrick Nolan), anti-HA.11 Epitope Tag antibody (1/1000, Clone 16B12, Biolegend), anti-ATBF1 (1/1000, AT-6 PD011, MBL) and anti-Flag M2 (1/2000, F3165-.2MG, Sigma-Aldrich). Blots were stripped twice. Imaging was performed with an Amersham Imager 600 CCD camera (GE Healthcare, Chicago, Illinois, USA).

**DNA methylation analysis**

Peripheral whole-blood DNA from ten patients within our LoF cohort (samples used for methylome analysis are highlighted in Supplemental Table 7) was extracted in different institutions using standard techniques and bisulfite conversion followed to pursue the DNA methylation experiment. DNA methylation analysis of the samples was performed using the Illumina Infinium methylation EPIC bead chip arrays according to the manufacturer's protocol. These arrays cover above 860,000 human genomic methylation CpG sites, including 99% of RefSeq genes and 96% of CpG islands. The resulting methylated and unmethylated signal intensity data were imported into R 4.1.2 for analysis. Normalization was performed by the Illumina normalization method with background correction using the minfi package[77]. Probes located on X and Y chromosomes, known to cross-react with chromosomal locations other than their target regions, known to contain SNPs at or near CpG interrogation or single nucleotide extension sites, or suggested by Illumina to be problematic were excluded. Arrays with more than 5% probe failure rate were eliminated from the analysis. Samples were examined for genome-wide methylation density, and those deviating from a bimodal distribution were excluded (all samples passed). Principal component analysis (PCA) was applied to observe the overall structure of the batches and to identify outliers.

Using the MatchIt package[78], 50 control samples matched to the ZFHX3 samples by age, sex, and array type were selected from the EpiSign Knowledge Database (EKD, https://episign.lhsc.on.ca/index.html). For each probe, methylation level, called β-value, calculated as the ratio of methylated signal intensity over the sum of methylated and unmethylated signal intensities, was converted to M-values using logit transformation. Using the limma package[79], linear regression was performed, where blood cell type compositions, estimated by the Houseman's algorithm[80], were added to the model matrix. Using eBayes function in the limma package, the resulting p-values were moderated.

Probe selection was performed in three steps. First, 1000 probes with the highest product of mean methylation difference between the ZFHX3 case samples and control samples and the negative of the logarithm of p-values were selected. Next, 333 probes with the highest areas under the receiver's operating characteristic (ROC) curve were retained. Finally, probes with pairwise Pearson's correlation coefficients >0.75 were eliminated, resulting in 208 probes.

Using the selected set of probes, hierarchical clustering by Ward's method on Euclidean distance and multidimensional scaling (MDS) by scaling of the pairwise Euclidean distances between samples were performed.

The 208 probes were then used for constructing two support vector machine (SVM) classifiers using the e1071 package, to generate methylation variant pathogenicity (MVP) scores ranging from 0 to 1 for each sample, where scores near 1 indicate similarity to the identified methylation profile for the *ZFHX3* haploinsufficiency. The first classifier was constructed by training the 10 ZFHX3 case samples only against the 50 matched control samples, and other control samples and samples from other disorders with known episignatures from EKD were added to the model for assessing the specificity. The second classifier was constructed by training the 10 ZFHX3 samples against the matched control samples and 75% of other control samples and samples with other disorders from the EKD, and the remaining 25% were used for testing. Three DNA samples from patients with ZFHX3 missense variants (Supplementary Table 8) were also tested for the methylation profile of the LoF cohort. A cohort of subjects with BAFopathy from the EKD were also supplied into the model in order to observe any overlap between the methylation profiles of ZFHX3 haploinsufficiency and BAFopathy.

**ChIP-sequencing and data analysis**

Chromatin was sheared using Bioruptor® sonication device (Diagenode Cat# B01020001) combined with the Bioruptor® Water cooler for 20 using a 30'' [ON] 30'' [OFF] settings for a total of 20 minutes, the most effective shearing time. Shearing was performed in 1,5 ml Bioruptor® Microtubes (Diagenode Cat# C30010010-300) in ~ 150μl. An aliquot of this chromatin was used to assess the size of the DNA fragments obtained by High Sensitivity NGS Fragment Analysis Kit (DNF-474) on a Fragment Analyzer™ (Agilent).

ChIP was performed manually following the protocol of the aforementioned kit. Chromatin corresponding to 5μg was immunoprecipitated using 3μg for the following antibody, anti-ZFHX3 (custom antibody (Parsons et al., 2015)). Chromatin corresponding to 1% was set apart as Input. qPCR analyses were made to check ChIP efficiency using KAPA SYBR® FAST

(Sigma-Aldrich) on LightCycler® 96 System (Roche) and results were expressed as % recovery = 2^(Ct_input-Ct_sample).

The library preparation has been conducted by Diagenode ChIP-seq/ChIP-qPCR Profiling service (Diagenode Cat# G02010000). Libraries were prepared using IP-Star® Compact Automated System (Diagenode Cat# B03000002) from input and ChIP'd DNA using MicroPlex Library Preparation Kit v3 /96 rxns (Diagenode Cat# C05010002) with 24 UDI for MicroPlex v3 - Set I and Set II (Diagenode Cat# C05010008 and C05010009). Optimal library amplification was assessed by qPCR using KAPA SYBR® FAST (Sigma-Aldrich) on LightCycler® 96 System (Roche) and by using High Sensitivity NGS Fragment Analysis Kit (DNF-474) on a Fragment Analyzer™ (Agilent). Libraries were then purified using Agencourt® AMPure® XP (Beckman Coulter) and quantified using Qubit™ dsDNA HS Assay Kit (Thermo Fisher Scientific, Q32854). Finally their fragment size was analised by High Sensitivity NGS Fragment Analysis Kit (DNF-474) on a Fragment Analyzer™ (Agilent). Libraries were pooled and sequenced on a Illumina Novaseq 6000 (Novaseq Control Software 1.7.0) with paired-end reads of 50bp length. Trimming of the adaptors was performed with cutadapt, followed by a quality control of the trimmed reads using FastQC[81]. Trimmed reads were aligned to the reference genome (hg38) using BWA software v.0.7.17[82]. Samples were filtered for regions blacklisted by the ENCODE project[83]. PCR duplicates and multimapping reads were removed using samtools[84]. Alignment coordinates were converted to BED format using BEDTools v.2.17[85] and peak calling was performed using epic2 with optimized parameters for transcription factors[86]. The peak calling scores were normalized by dividing each peak score with the maximum peak score value of each set. The intersect of the peaks called for endogenous and ectopic ZFHX3 ChIP-seq was determined via BEDtools intersect.

These intersect peaks were annotated using ChIPSeeker as mentioned in the main manuscript.

#### *Drosophila* stock and the generation of the transgenic lines

The following fly lines used in this study were obtained from the Bloomington *Drosophila* Stock Center (BDSC), Vienna *Drosophila* Resource Center (VDRC) or generated in house or by Genetivision. A wildtype control line used for the *Drosophila* experiments is $w^{1118}$ from BDSC. The zfh2 lines are:

- y[1] v[1]; P{y[+t7.7] v[+t1.8]=TRiP.HMC03043}attP2 (50643, BDSC) = TRIP-*Zfh2* RNAi

- w; UAS-Zfh2:RNAi/SM6A-TM6B/UAS-Zfh2:RNAi (provided by Prof. Fernando J Díaz-Benjumea, Centro de Biología Molecular Severo Ochoa, UAM)
- UAS-zfh2 RNAi/TM6B (13305, VDRC)
- y[1] w[*]; TI{GFP[3xP3.cLa]=CRIMIC.TG4.2}zfh2[CR01648-TG4.2]/In(4)ci[D]ci[D] pan[ciD] (86479, BDSC) = *Zfh2*-CRIMIC/GFP
- *Zfh2*-CRIMIC/GFP; UAS-RFP (generated in house)
- w[1118]; PBac{y[+mDint2] w[+mC]=zfh2-GFP.FPTB}VK00033/TM3, Sb[1] w[*] (38674, BDSC)

The ZFHX3 fly lines are:

- Puast attP2 ZFHX3/TM3 m9m1 (Genetivision)
- Puast attP40 ZFHX3/CyO m1m1 (Genetivision)

#### Immunofluorescence and confocal microscopy for larval brains

Larval brains from the fly lines $w^{1118}$ (#, BDSC), CRIMIC/GFP;UAS-RFP and PBac{y[+mDint2] w[+mC]=zfh2-GFP.FPTB}VK00033/TM3, Sb[1], w[*] (38674, BDSC were dissected in PBS, and fixed in 4% paraformaldehyde in PBS, for 10 min at room temperature. The samples were then washed in 1× PBT (PBS+ 0.2% Triton X-100) and blocked with 10% normal goat serum in 0.5% PBS titon X-100 (PBST). The primary antibodies used for the endogenous zfh2 and zfh2 GFP-tagged expression detection is the rat Zfh2 antibody (1:400)[87], kindly provided by the lab of Chris Doe, University of Oregon and a mouse GFP antibody (catalog number, company). The secondary antibodies used were goat anti-rat IgG (A48264) and anti-mouse (A21203) at a 1:1,000 dilution, conjugated to Alexa 594 and Alexa 488, respectively. The samples were washed three times for 5 min with 0.5% PBST. From the CRIMIC/GFP; UAS-RFP line, larval brains were dissected in PBS, and fixed in 4% paraformaldehyde in PBS, for 10 min at room temperature and no antibodies were use. Furthermore, all larval brains are mounted on a glass slide using Fluoromount-G™ Mounting Medium, with DAPI (Thermofisher Scientific). The imaging was performed with a Leica SP8 confocal microscope and processed using ImageJ.

### Supplemental Figures

**Supplemental Figure 1.** Rows 1-2: Front and side profile pictures of probands with a microdeletion containing ZFHX3. Rows 3-5: Front and side profile pictures of probands with a ZFHX3 protein truncating variant (PTV). Row 6: Front and side profile picture of probands with a multigenic deletions containing ZFHX3. Rows 7-8: Hands and digits of probands with a microdeletion (row 7) or PTV (row 8) affecting ZFHX3.

**Supplemental Figure 2.** ZFHX3 expression on the RNA level A. in 54 non-diseased adult tissues (GTEx data v8; downloaded from https://gtexportal.org/home/gene/ZFHX3), B. in 22 developmental stages across 7 human tissues (downloaded from https://apps.kaessmannlab.org/evodevoapp/), C. during neuronal differentation from pluripotent stem cells towards neural progenitor cells (NPCs) and mature neurons (single cell expression data downloaded from https://bioinf.eva.mpg.de/shiny/sample-apps/scApeX/.

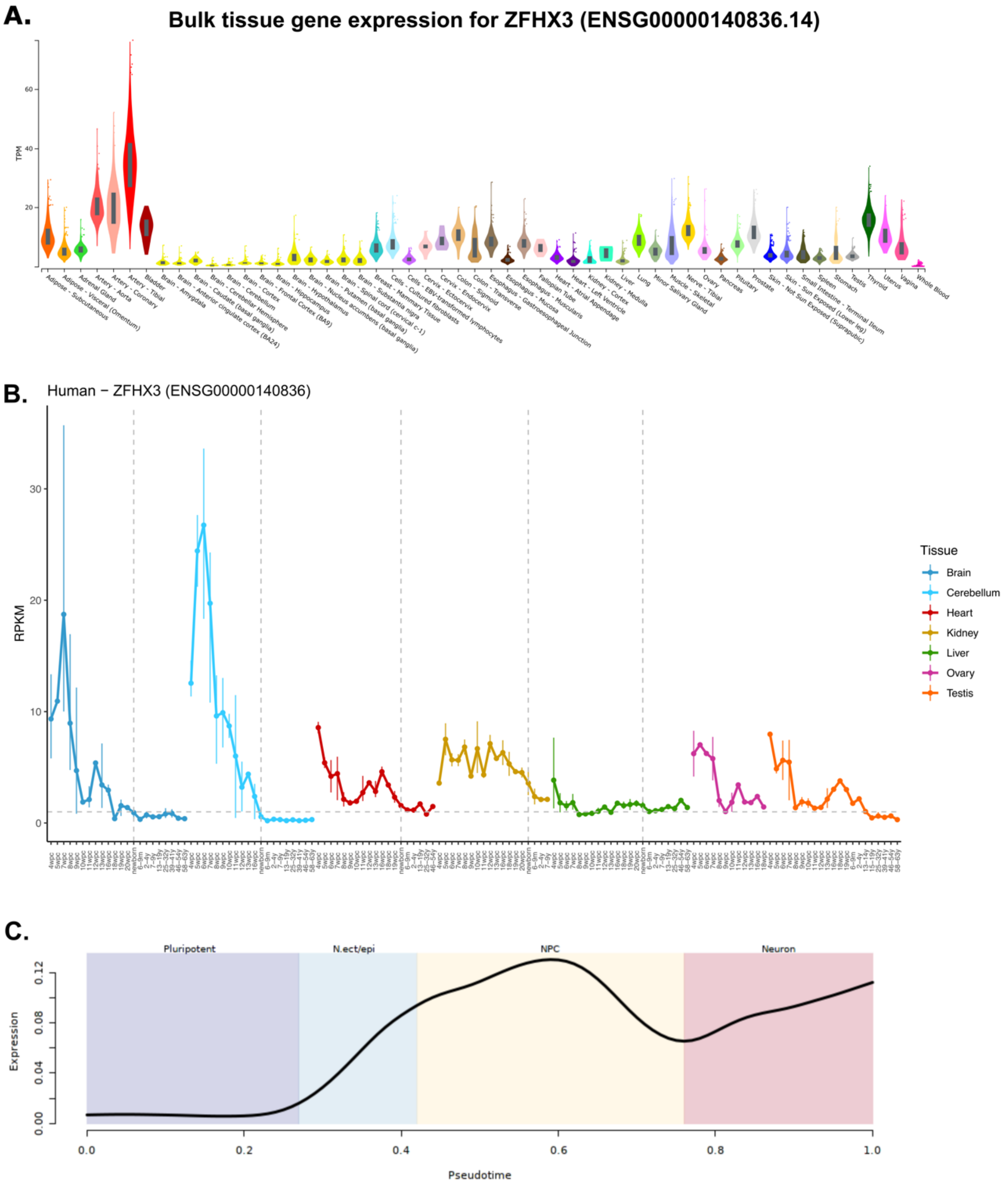
A.
Bulk tissue gene expression for ZFHX3 (ENSG00000140836.14)
TPM
B.
Human − ZFHX3 (ENSG00000140836)
RPKM
Tissue
Brain
Cerebellum
Heart
Kidney
Liver
Ovary
Testis
C.
Pluripotent
N.ect/epi
NPC
Neuron
Expression
Pseudotime

**Supplemental Figure 3.** ZFHX3 expression in single-cell sequencing data generated from single neural rosettes (SNR) derived organoids by the Shcheglovitov lab in their online browser http://organoid.chpc.utah.edu. In the dot plot, the color indicates the average expression of ZFHX3 per cluster and the size of the dot indicates the percentage of cells within the cluster expressing ZFHX3. *ZFHX3* is particularly expressed in 1 month old organoids and in inhibitory neurons, less in 5-month-old organoids.

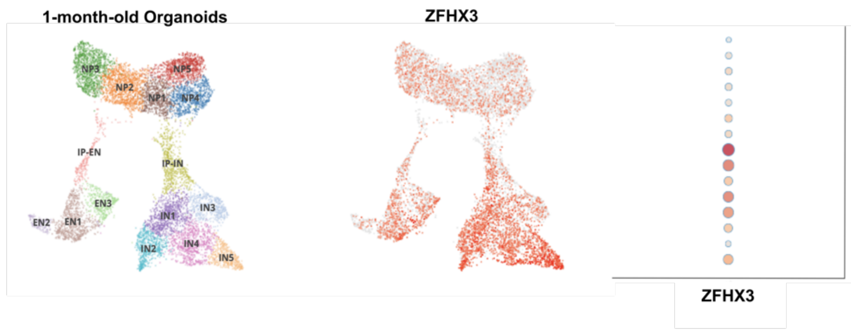


| | Average Expression | % cells |
|---|---|---|
| NP5 | 0,171 | 15,62 |
| NP4 | 0,3 | 26,53 |
| NP3 | 0,405 | 34,75 |
| NP2 | 0,346 | 30,27 |
| NP1 | 0,296 | 26,61 |
| IP-IN | 0,568 | 41,71 |
| IP-EN | 0,397 | 32,17 |
| IN5 | 1,721 | 90,93 |
| IN4 | 1,218 | 75,29 |
| IN3 | 0,594 | 43,66 |
| IN2 | 1,173 | 71,26 |
| IN1 | 1,021 | 66,12 |
| EN3 | 0,587 | 44,67 |
| EN2 | 0,255 | 21,94 |
| EN1 | 0,802 | 58,32 |

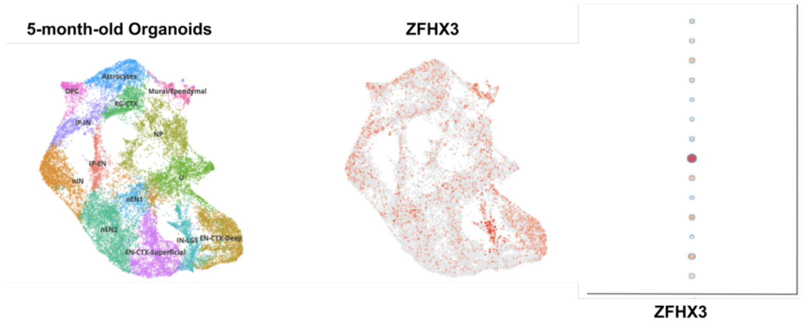


| | Average Expression | % cells |
|---|---|---|
| U | 0,087 | 15,62 |
| RG-CTX | 0,065 | 26,53 |
| OPC | 0,117 | 34,75 |
| NP | 0,074 | 30,27 |
| nIN | 0,0630 | 26,61 |
| nEN2 | 0,055 | 41,71 |
| nEN1 | 0,070 | 32,17 |
| Mural/Ependymal | 0,275 | 90,93 |
| IP-IN | 0,110 | 75,29 |
| IP-EN | 0,050 | 43,66 |
| IN-LGE | 0,148 | 71,26 |
| EN-CTX-Superficial | 0,043 | 5,96 |
| EN-CTX-Deep | 0,138 | 18,82 |
| Astrocytes | 0,087 | 11,78 |

NP = neural progenitor clusters, IP-IN = intermediate neural progenitor for inhibitory neurons, IP-EN = intermediate neural progenitor for excitatory neurons, (n)IN = newborn inhibitory neurons, (n)EN = newborn excitatory neurons, U= unknown, RG-CTX = radial glia of the cortex, OPC= oligodendrocyte progenitor cell, LGE= lateral ganglionic eminence.

**Supplemental Figure 4.** According to GTEx v8, ENST00000397992.5 (short isoform) is more abundantly expressed in adult tissues in comparison with ENST000002694929.10 (long isoform). Figure downloaded from https://gtexportal.org/home/gene/ZFHX3 (isoform detection). However, it has to be noted that both isoforms can only be distinguished by the presence of exon 1-2 of the long isoform.

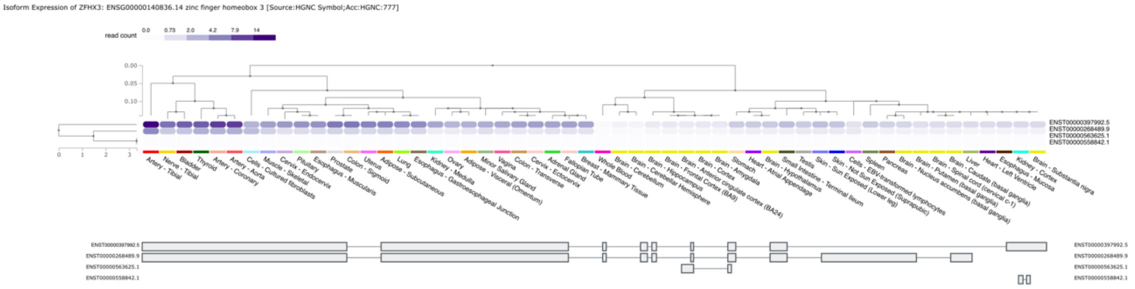


**Supplemental Figure 5. No co-localization with nestin, a known neural cytoplasmic marker, was observed in NSCs**, thereby demonstrating that in NSCs, ZFHX3 is only present in the nucleus. Images were merged to observe the contrast between the compartments. Scale bar: 10µm.; hESC: human embryonic stem cells; NSC: neural stem cells.

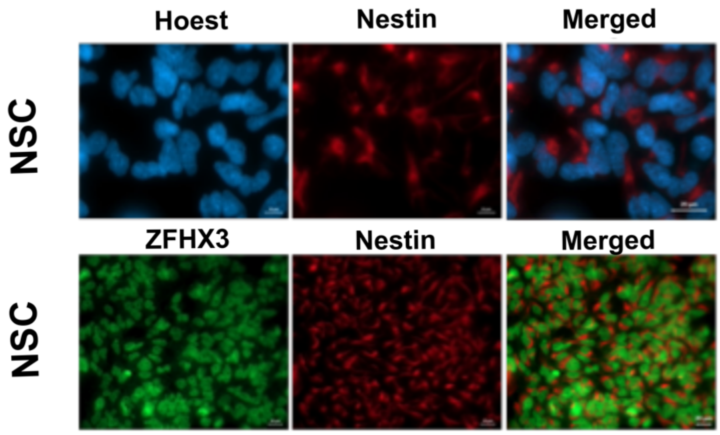


**Supplemental Figure 6:** Metascape enrichment of the ZFHX3 interactome. Shown are heatmaps of enriched GO-terms across input protein lists (= ZFHX3 and 57 interactors), colored by p-values.

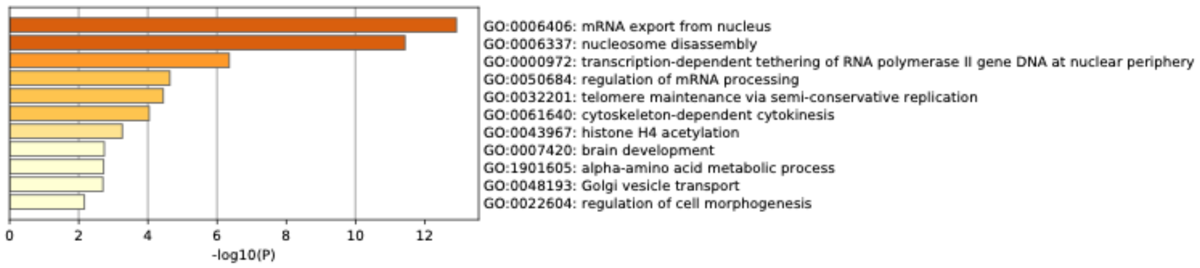

**Supplemental Figure 7. A.** Protein expression of SMARCB1-3xHA and CPSF2-3xHA after co-transfection in HEK293T cells. **B.** Protein expression of ZFHX3-Flag, detected with anti-Flag and anti-ATBF1, after co-transfection.

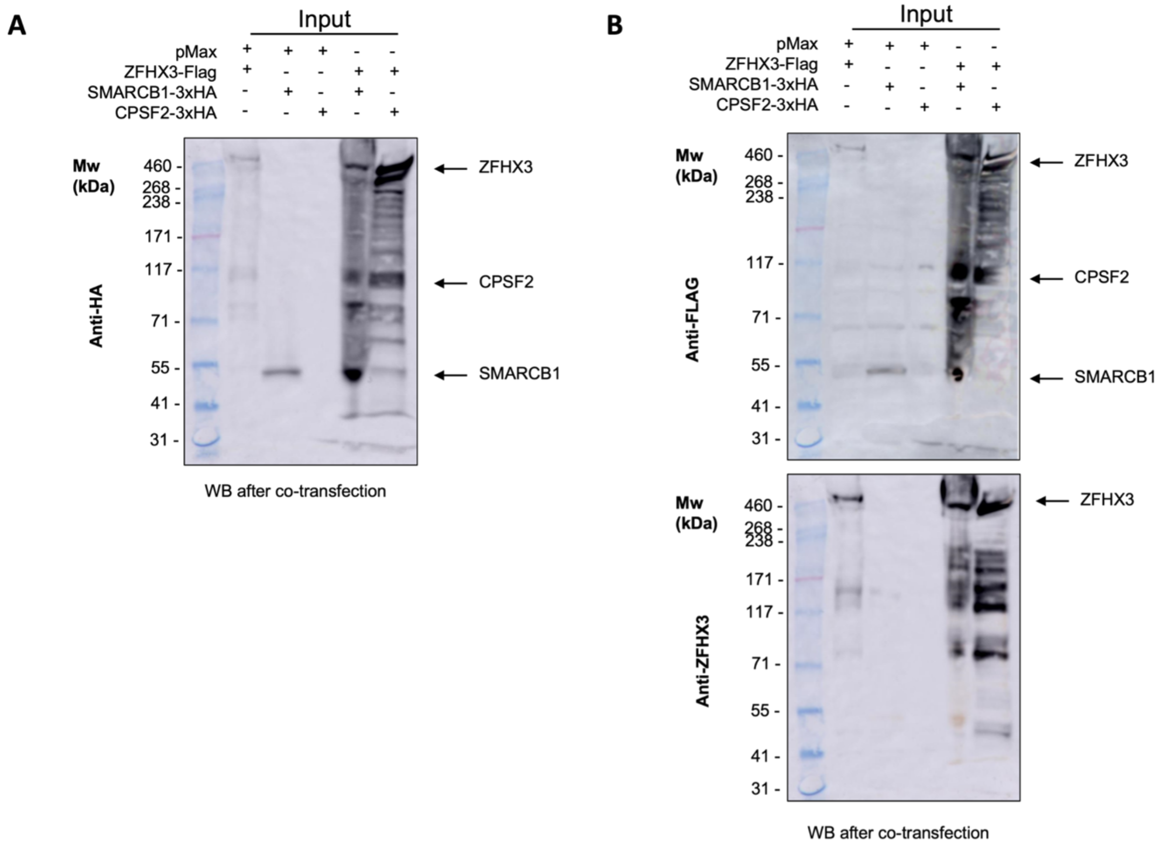


**Supplemental Figure 8. A.** Vector map of pcDNA3-FLAG-ATBF1 (#40927, AddGene) co-transfected in HEK293T cells followed by co-immunoprecipitation. **B.** Vector map of a newly designed pMH-HA N-term tag expression vector with the Open Reading Frame (ORF) of SMARCB1 (left) and CPSF2 (right) via Gateway cloning (Gateway Technology, Gateway LR reaction, Invitrogen), respectively co-transfected in HEK293T cells followed by co-immunoprecipitation. SMARCB1 and CPSF2 constructs were ordered from Genecopoeia as shuttle vectors.

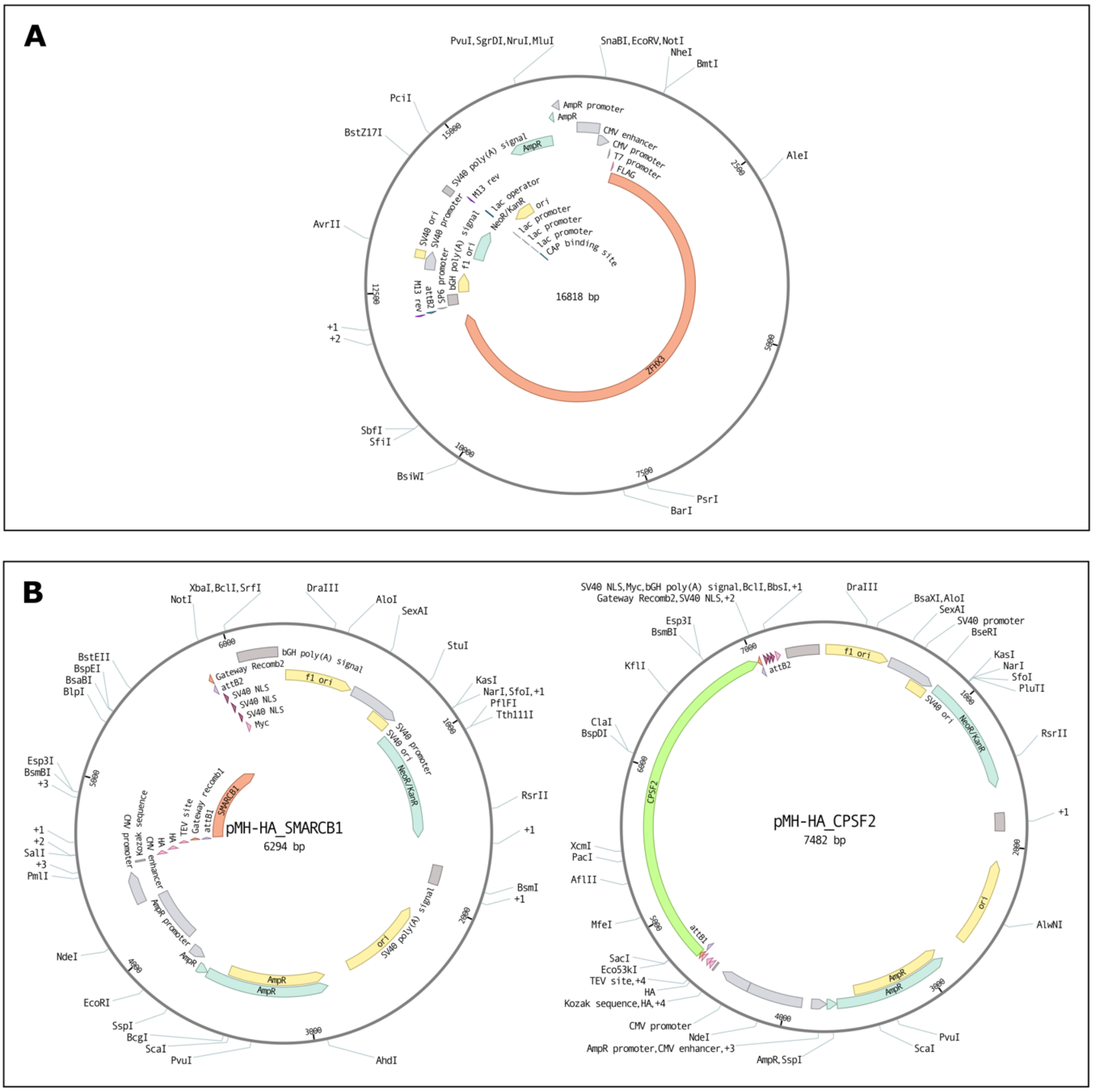
A
PvuI,SgrDI,NruI,MluI
SnaBI,EcoRV,NotI
NheI
BmtI
PciI
BstZ17I
AleI
AvrII
+1
+2
SbfI
SfiI
BsiWI
PsrI
BarI
AmpR promoter
AmpR
CMV enhancer
CMV promoter
T7 promoter
FLAG
SV40 poly(A) signal
M13 rev
lac operator
ori
NeoR/KanR
lac promoter
lac promoter
CAP binding site
SV40 ori
SV40 promoter
bGH poly(A) signal
f1 ori
SP6 promoter
attB2
M13 rev
ZFHX3
16818 bp
15000
2500
12500
5000
10000
7500
B
XbaI,BclI,SrfI
NotI
DraIII
AloI
SexAI
StuI
KasI
NarI,SfoI,+1
PflFI
Tth111I
BstEII
BspEI
BsaBI
BlpI
Esp3I
BsmBI
+3
+1
+2
SalI
+3
PmlI
RsrII
+1
BsmI
+1
NdeI
EcoRI
SspI
BcgI
ScaI
PvuI
AhdI
bGH poly(A) signal
Gateway Recomb2
attB2
SV40 NLS
SV40 NLS
SV40 NLS
Myc
f1 ori
SV40 promoter
SV40 ori
NeoR/KanR
SV40 poly(A) signal
ori
AmpR
AmpR
AmpR
AmpR promoter
CMV enhancer
CMV promoter
Kozak sequence
HA
HA
TEV site
Gateway recomb1
attB1
SMARCB1
pMH-HA_SMARCB1
6294 bp
SV40 NLS,Myc,bGH poly(A) signal,BclI,BbsI,+1
Gateway Recomb2,SV40 NLS,+2
DraIII
BsaXI,AloI
SexAI
SV40 promoter
BseRI
Esp3I
BsmBI
KflI
KasI
NarI
SfoI
PluTI
RsrII
ClaI
BspDI
+1
XcmI
PacI
AflII
AlwNI
MfeI
SacI
Eco53kI
TEV site,+4
HA
Kozak sequence,HA,+4
CMV promoter
NdeI
AmpR promoter,CMV enhancer,+3
AmpR,SspI
ScaI
PvuI
attB2
f1 ori
SV40 ori
NeoR/KanR
CPSF2
attB1
ori
AmpR
AmpR
pMH-HA_CPSF2
7482 bp

**Supplemental Figure 9. Expression of zfh2 (RFP) and Elav (GFP) in the third instar larval brain in wild-type line**. Elav is shown in green, zfh2 is shown in red and DAPI is shown in dark blue. The immunofluorescence performed on the third instar larval brains of WIII line localizes zfh2 to the nucleus of the neurons (elav, nuclear neuronal marker).

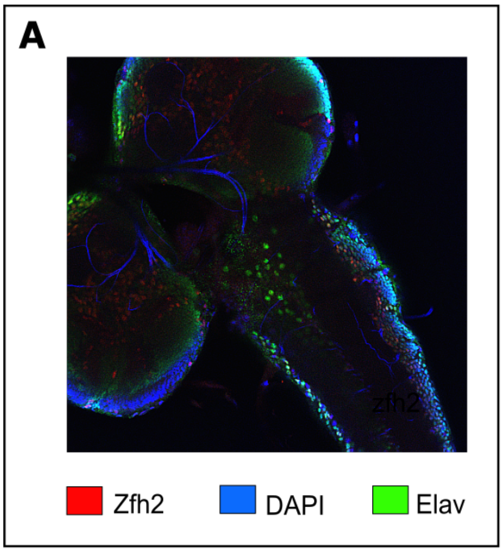


### Supplemental Tables

**Supplemental Table 1.** Clinical characteristics of seventeen individuals and two affected family members with a microdeletion affecting ZFHX3 (probands 1-18). Individuals for whom the clinical characteristics were not taken into account for the general phenotype are highlighted in yellow. Individuals for whom we only have limited clinical information, are highlighted in light red.

**Supplemental Table 2.** Clinical characteristics of twenty-three individuals and four affected family members with a PTV affecting ZFHX3 (probands 19-42).

**Supplemental Table 3.** Missense variants in gnomAD v3.1.1 affecting the same codon as the truncating variant in probands 21, 36, 37 and 42

**Supplemental Table 4.** Spearman correlation values of gene expression values compared to ZFHX3 expression in 7 human tissues during 22 developmental stages (Cardoso-Moreira et al., 2019). Only genes for which the rho value was significant (p<=0.05) are given.

**Supplemental Table 5.** IP-MS results for ZFHX3 in SH-SY5Y. ZFHX3 is highlighted in blue and appears one of the top enriched proteins. Proteins highlighted in green are significantly enriched (FDR <=0.05) compared to the IgG control. Proteins highlighted in red are significantly enriched (FDR <=0.05) in the IgG control.

**Supplemental Table 6.** IP-MS results for ZFHX3 in NSC. ZFHX3 is highlighted in blue and appears one of the top enriched proteins. Proteins highlighted in green are significantly enriched (FDR <=0.05) compared to the IgG control.Proteins highlighted in red are significantly enriched (FDR <=0.05) in the IgG control.

**Supplemental Table 7.** *ZFHX3* loss-of-function patients' samples list for methylome analysis

**Supplemental Table 8.** *ZFHX3* missense patients' samples list for methylome analysis in probands A, B and C

**Supplemental Table 9.** Other protein coding genes residing in the CNVs of Probands 1-18

**Supplementary Table 10**. RT-qPCR primers used expression profiling and PCR primers used to test the genotype of the *Drosophila* lines.

**Supplementary Table 11**. The intersect peaks between endogenous and ectopic ZFHX3 ChIP-sequencing.
